# Second-line tenofovir alafenamide for children with HIV in Africa

**DOI:** 10.1101/2024.04.12.24304337

**Authors:** Victor Musiime, Alexander J Szubert, Hilda A Mujuru, Cissy Kityo, Katja Doerholt, Shafic Makumbi, Veronica Mulenga, Wedu Ndebele, Mwate Mwamabazi, Helen McIlleron, Mutsa Bwakura-Dangarembizi, Eva Natukunda, Kyomuhendo Jovia Linda, Lara Monkiewicz, Monica Kapasa, Mary Nyathi, Bwendo Nduna, Annabelle South, Godfrey Musoro, Khozya Zyambo, Yingying Zhang, Simon Walker, Anna Turkova, A Sarah Walker, Alasdair Bamford, Diana M Gibb, CHAPAS-4 Trial Team

## Abstract

**Background:** Children living with HIV have few second-line antiretroviral therapy(ART) options, especially fixed-dose-combinations(FDC).

**Methods:** Children from Uganda, Zambia, Zimbabwe were randomised to second-line tenofovir alafenamide(TAF)/emtricitabine(FTC) or standard-of-care(SOC) backbone (abacavir(ABC) or zidovudine(ZDV) with lamivudine(3TC)) in the factorial CHAPAS-4 trial. The second randomisation (reported elsewhere) was to dolutegravir(DTG), ritonavir-boosted darunavir(DRV/r), atazanavir(ATV/r) or lopinavir(LPV/r) as anchor drug. All drugs were dosed using WHO weight-bands and children <25kg received a new paediatric TAF/FTC(15/120mg) FDC tablet. The primary endpoint was viral load(VL)<400copies/ml at week-96, analysed using logistic regression, hypothesising that TAF/FTC would be non-inferior to SOC (10% margin). Secondary endpoints included safety and immunological outcomes. Analyses were intention-to-treat.

**Results:** 919 children 3–15years, 497(54%) male, median[IQR] baseline viral load(VL) 17,573copies/ml [5549-55,700] and CD4 count 669cells/mm^3^[413-971], spent 99% of time on allocated NRTI backbone. At week-96, 406/454(89.4%) receiving TAF/FTC vs. 378/454(83.3%) receiving SOC had VL<400copies/mL (adjusted difference[95%CI]: 6.3%[2.0%,10.6%], p=0.004), with no evidence that this varied by ABC/3TC or ZDV/3TC SOC. CD4 count improved similarly in both arms. Growth was better with TAF/FTC vs. SOC, without evidence of excess weight-gain with any backbone/anchor drug combination (including DTG±TAF/FTC, interaction p=0.51). Bone health parameters were similar between arms, irrespective of anchor drug. One child died (treatment-unrelated); 29(3%) had serious adverse events without differences between arms.

**Conclusions:** TAF/FTC was virologically superior to SOC ZDV/3TC or ABC/3TC with a favourable safety profile, irrespective of anchor drug. Development of child-friendly TAF/FTC FDCs (±anchor drug) would increase cost-effective ART options for children and reduce drug access gaps between children and adults.(ISRCTN22964075)

## Background

At the end of 2022, of the estimated 1.5 million children living with HIV (CLHIV) under 15 years globally, 57% were receiving antiretroviral therapy (ART), the majority living in Africa.^1-3^

Estimates from global cohort studies suggest that around 5-10% children have switched to second-line ART,^4,5^ but few to third-line. The need for access to safe, effective ART options for children with first-line ART regimen failure will increase in coming years, even as dolutegravir (DTG) is rolled-out for children for both first-and second-line ART.

The World Health Organization (WHO) currently recommends two nucleoside/nucleotide reverse transcriptase inhibitor drugs (NRTIs) as a backbone in both first-line and second-line ART regimens. These are sequenced between abacavir/lamivudine (ABC/3TC) and zidovudine/lamivudine (ZDV/3TC), from first-line to second-line ART. Tenofovir disoproxil fumarate (TDF) with 3TC or emtricitabine (FTC) is recommended for first and/or second-line ART for adolescents >30kg. Data from adult trials suggests tenofovir is superior to ZDV.^6,7^ However, concerns about bone health, renal toxicity and poor availability of paediatric formulations limits use of TDF in younger children.^8^

Tenofovir alafenamide (TAF)/emtricitabine (FTC) (15/120mg) is a new small, paediatric fixed-dose-combination (FDC) including TAF, a prodrug of tenofovir, with lower milligram dosage and more favourable bone and renal safety profiles than the TDF prodrug. Unlike TDF, where tenofovir is widely distributed following absorption, tenofovir from TAF remains mostly restricted to cells with high carboxyesterase and catepsin A activity such as hepatocytes and lymphocytes, resulting in lower tenofovir plasma levels but higher intracellular concentrations and lower toxicity compared to TDF.^9,10^ There are limited data on TAF in children in Africa. We recently reported the first pharmacokinetic (PK) data for African children aged 3-15 years in CHAPAS-4 receiving TAF/FTC with either DTG or a boosted protease inhibitor (bPI), showing that tenofovir concentrations were similar to those that are safe and effective in adults.^11^

There are no paediatric data comparing TAF/FTC to WHO-recommended NRTI backbone options (ABC/3TC and ZDV/3TC). In CHAPAS-4, we compared the efficacy and safety of TAF/FTC vs. ABC/3TC or ZDV/3TC in African children starting second-line ART.

## Methods

CHAPAS-4 (ISRCTN22964075) was a randomised, open-label trial with a factorial design (2X4). Children were randomised to one of two NRTI backbones (TAF/FTC or SOC (ABC/3TC or ZDV/3TC, whichever had not been used first-line)). They were simultaneously randomized to one of four anchor drugs (DTG, darunavir/ritonavir (DRV/r), atazanavir/ritonavir (ATV/r) or lopinavir/ritonavir (LPV/r), reported elsewhere). The doses of trial drugs followed WHO weight bands (Table S1). Of note, the TAF doses were the same irrespective of anchor drug.

The trial was approved by ethics committees in Uganda, Zambia, Zimbabwe, South Africa and United Kingdom. The protocol is available at www.mrcctu.ucl.ac.uk/studies/all-studies/c/chapas-4. Participants were recruited at six hospitals in three sub-Saharan African countries: Uganda (Joint Clinical Research Centre, Kampala; Joint Clinical Research Centre, Mbarara), Zambia (University Teaching Hospital, Lusaka; Arthur Davison Children’s Hospital, Ndola) and Zimbabwe (University of Zimbabwe Clinical Research Centre, Harare; Mpilo Central Hospital, Bulawayo).

Participants were CLHIV aged 3-15 years, weighing ≥14kg, requiring switch from first-line NNRTI-based ART for virologic failure defined as viral load(VL)>1000 copies/ml with/without immunological and/or clinical failure. Children had to be able to swallow tablets, and post-menarchal females required a negative pregnancy test. Guardians provided written informed consent, with additional assent from older children, according to national guidelines. Children were excluded if they had severe hepatic impairment (alanine aminotransferase (ALT) ≥5 times upper-limit of normal (ULN), or ALT ≥3xULN and bilirubin ≥2xULN or clinical liver disease).

Randomisation was stratified by centre and first-line NRTI (ABC/3TC or ZDV/3TC). A computer-generated sequential randomisation list with variably sized permuted blocks was prepared by the trial statistician and incorporated securely into an online trial database. The allocation was concealed until eligibility was confirmed by local centre staff, who then performed the randomisation.

Participants were seen at screening, ART switch (week 0), 2, 6, 12 weeks and 12 weekly thereafter to at least 96 weeks (primary endpoint): extended follow-up continued through 2 February 2023. Visits included clinical and laboratory efficacy and safety assessments. Children with prevalent tuberculosis at enrolment or subsequent incident tuberculosis had TAF and anchor drugs changed during anti-tuberculosis treatment to adjust for rifampicin interaction. Measures were taken to ensure participant follow up during the COVID-19 pandemic (Panel 1 in Supplementary Appendix 1). Primary outcome was VL <400 copies/ml at week 96 (death counted as ≥400). Secondary efficacy outcomes were VL <60 and <1000 copies/ml at week 96, death/WHO 3/4 events, changes in CD4 (absolute and percentage), and genotypic resistance (assays ongoing). Secondary safety outcomes were grade 3/4, serious, and ART-modifying adverse events (AEs); and changes in cholesterol (total, low-density lipoprotein (LDL), high-density lipoprotein (HDL), triglycerides, bilirubin and creatinine clearance (CrCl). Other outcomes included changes in weight-, height- and body mass index (BMI)- for-age and bone mineral density Z-scores.

In addition, cost-effectiveness analyses were conducted to compare TAF/FTC versus SOC arms over the full 96 weeks of the trial. Health was measured in quality-adjusted life-years (QALYs) and costs, clinic visits and hospital stays, both discounted at 3% per annum. Further detail is included on cost effectiveness in Supplementary Appendix 2.

Assuming 80.0%-87.5% of children on SOC achieved VL <400 copies/ml at week 96, 920 children provided ≥95% power to demonstrate that TAF was non-inferior (10% margin) to SOC (two-sided alpha=5%), assuming 2.5% loss-to-follow-up (reduced from 10% in original protocol). An independent data monitoring committee reviewed the interim data at four meetings using the Haybittle–Peto criterion (99.9% confidence interval). Analyses were intention-to-treat. Analyses of the primary endpoint used logistic regression (adjusting for stratification factors), then marginal estimation of risk differences. Secondary per-protocol analysis included children who received the randomised NRTI backbone for >90% of follow-up. Sub-group analysis used interaction tests. For VL <60 and <1000 copies/ml, analysis was similar. For death/WHO 3/4 events, and grade 3/4, serious and ART-modifying AEs, groups were compared via Cox regression (unadjusted). Changes in continuous outcomes were analysed using Normal generalised estimating equations adjusting for visit, stratification factors and baseline (and interactions between these factors and visit), for an overall test of difference between groups over all visits (independent correlation). Analyses were conducted using Stata (version 17.0). The 95% confidence intervals were not adjusted for multiple testing.

The funder, European Developing Country Clinical Trial Partnership (EDCTP), and pharmaceutical companies donating additional funding (Gilead Sciences, Johnson and Johnson) and trial medications (ViiV Healthcare, Gilead sciences, Johnson and Johnson, CIPLA) did not participate in design, conduct or analysis of the trial.

## Results

919 children were randomised between 17 December 2018 and 1 April 2021, 458 to TAF/FTC and 461 to SOC (Figure 1). Baseline characteristics were similar between arms (Table 1 and Table S2). 497/919 (54.1%) children were male with median age 10 (inter-quartile range (IQR) 8,13) years. 777/919 (84.5%) were WHO stage 1/2. Median weight-for-age Z-score was -1.6 (IQR -2.4,-0.9); height-for-age Z-score -1.6 (−2.3,-0.8); body mass index (BMI)-for-age Z-score -1.0 (−1.7,-0.4). Median VL was 17573 copies/mL (IQR 5549,55700); CD4 count 669 cells/mm^3^ (413,971) and CD4% 28% (19%,36%). Median time on first-line ART was 5.6 years; prior to randomisation 44% were on nevirapine and 56% on efavirenz.

**Table 1:**
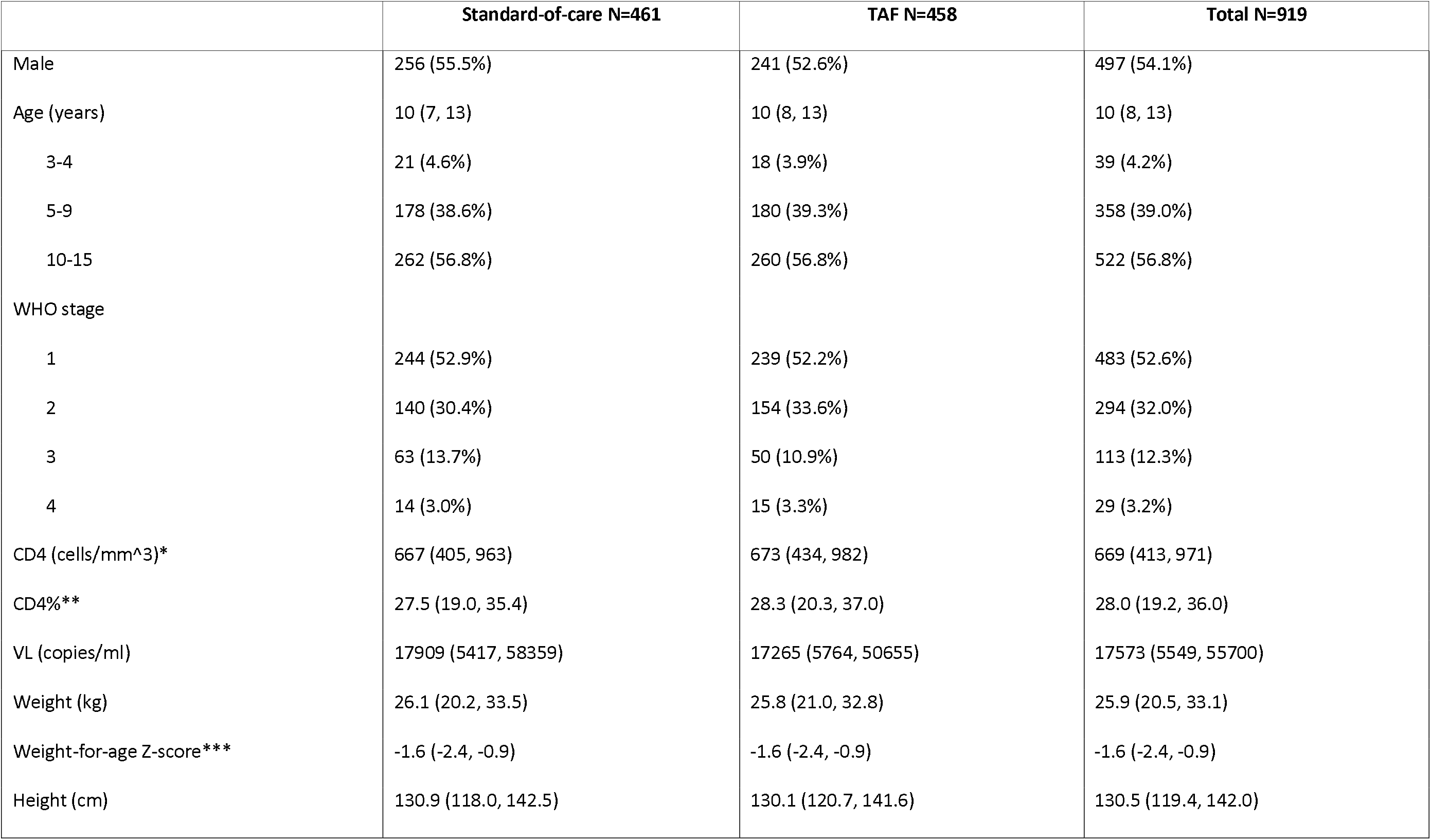
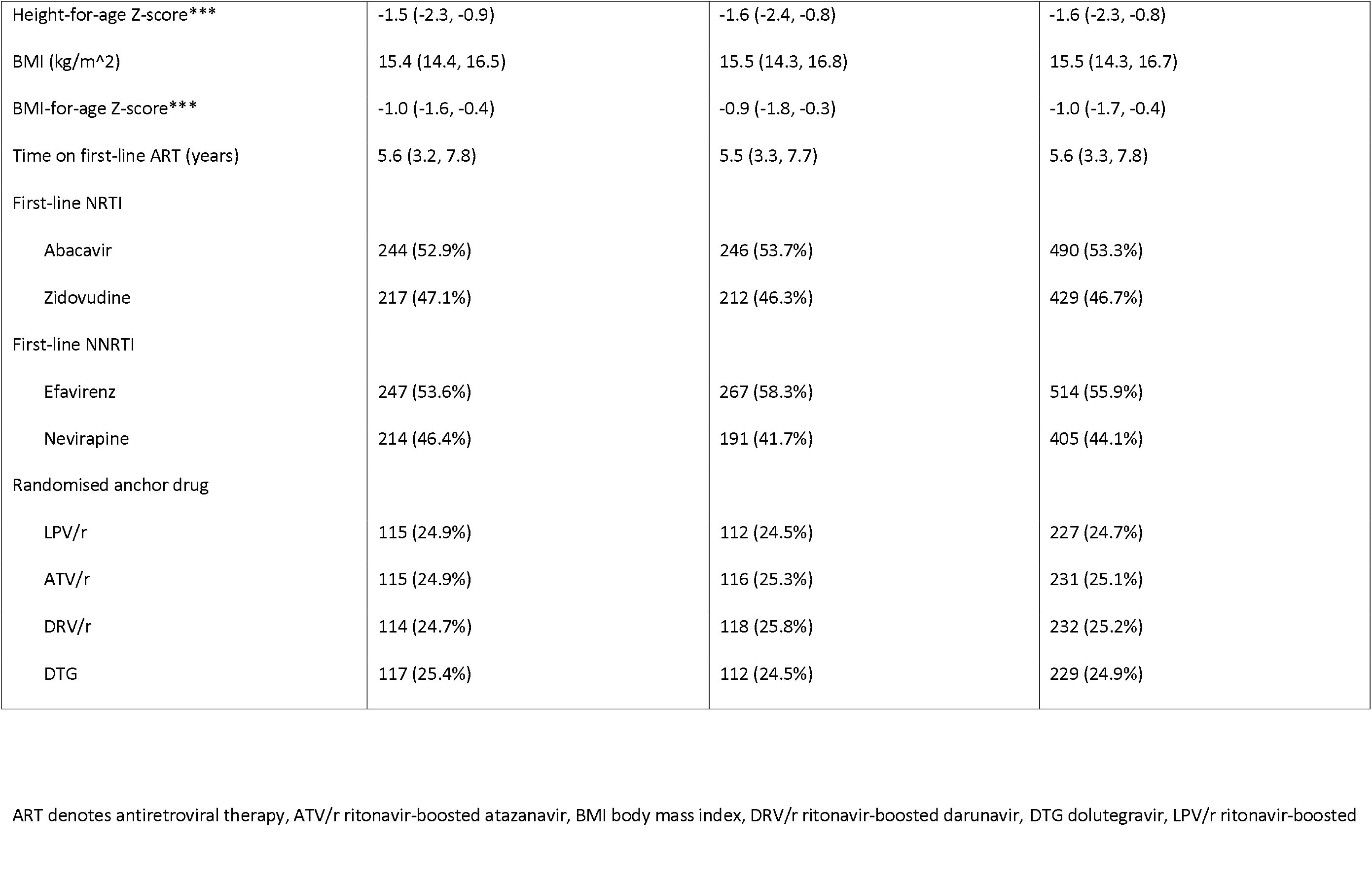
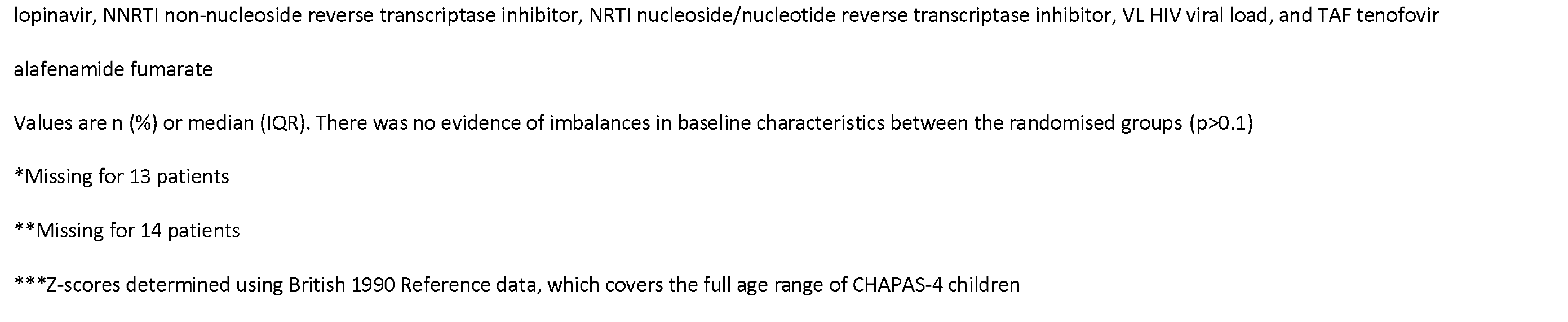
Baseline characteristics.

|  | Standard-of-care N=461 | TAF N=458 | Total N=919 |
| --- | --- | --- | --- |
| Male | 256 (55.5%) | 241 (52.6%) | 497 (54.1%) |
| Age (years) | 10 (7, 13) | 10 (8, 13) | 10 (8, 13) |
| 3-4 | 21 (4.6%) | 18 (3.9%) | 39 (4.2%) |
| 5-9 | 178 (38.6%) | 180 (39.3%) | 358 (39.0%) |
| 10-15 | 262 (56.8%) | 260 (56.8%) | 522 (56.8%) |
| WHO stage |  |  |  |
| 1 | 244 (52.9%) | 239 (52.2%) | 483 (52.6%) |
| 2 | 140 (30.4%) | 154 (33.6%) | 294 (32.0%) |
| 3 | 63 (13.7%) | 50 (10.9%) | 113 (12.3%) |
| 4 | 14 (3.0%) | 15 (3.3%) | 29 (3.2%) |
| CD4 (cells/mm<sup>3</sup>)* | 667 (405, 963) | 673 (434, 982) | 669 (413, 971) |
| CD4%** | 27.5 (19.0, 35.4) | 28.3 (20.3, 37.0) | 28.0 (19.2, 36.0) |
| VL (copies/ml) | 17909 (5417, 58359) | 17265 (5764, 50655) | 17573 (5549, 55700) |
| Weight (kg) | 26.1 (20.2, 33.5) | 25.8 (21.0, 32.8) | 25.9 (20.5, 33.1) |
| Weight-for-age Z-score*** | -1.6 (-2.4, -0.9) | -1.6 (-2.4, -0.9) | -1.6 (-2.4, -0.9) |
| Height (cm) | 130.9 (118.0, 142.5) | 130.1 (120.7, 141.6) | 130.5 (119.4, 142.0) |
| Height-for-age Z-score*** | -1.5 (-2.3, -0.9) | -1.6 (-2.4, -0.8) | -1.6 (-2.3, -0.8) |
| BMI (kg/m^2) | 15.4 (14.4, 16.5) | 15.5 (14.3, 16.8) | 15.5 (14.3, 16.7) |
| BMI-for-age Z-score*** | -1.0 (-1.6, -0.4) | -0.9 (-1.8, -0.3) | -1.0 (-1.7, -0.4) |
| Time on first-line ART (years) | 5.6 (3.2, 7.8) | 5.5 (3.3, 7.7) | 5.6 (3.3, 7.8) |
| First-line NRTI |  |  |  |
| Abacavir | 244 (52.9%) | 246 (53.7%) | 490 (53.3%) |
| Zidovudine | 217 (47.1%) | 212 (46.3%) | 429 (46.7%) |
| First-line NNRTI |  |  |  |
| Efavirenz | 247 (53.6%) | 267 (58.3%) | 514 (55.9%) |
| Nevirapine | 214 (46.4%) | 191 (41.7%) | 405 (44.1%) |
| Randomised anchor drug |  |  |  |
| LPV/r | 115 (24.9%) | 112 (24.5%) | 227 (24.7%) |
| ATV/r | 115 (24.9%) | 116 (25.3%) | 231 (25.1%) |
| DRV/r | 114 (24.7%) | 118 (25.8%) | 232 (25.2%) |
| DTG | 117 (25.4%) | 112 (24.5%) | 229 (24.9%) |

ART denotes antiretroviral therapy, ATV/r ritonavir-boosted atazanavir, BMI body mass index, DRV/r ritonavir-boosted darunavir, DTG dolutegravir, LPV/r ritonavir-boosted
lopinavir, NNRTI non-nucleoside reverse transcriptase inhibitor, NRTI nucleoside/nucleotide reverse transcriptase inhibitor, VL HIV viral load, and TAF tenofovir
alafenamide fumarate
Values are n (%) or median (IQR). There was no evidence of imbalances in baseline characteristics between the randomised groups ( $p>0.1$ )
\*Missing for 13 patients
\*\*Missing for 14 patients
\*\*\*Z-scores determined using British 1990 Reference data, which covers the full age range of CHAPAS-4 children

**Figure 1.**
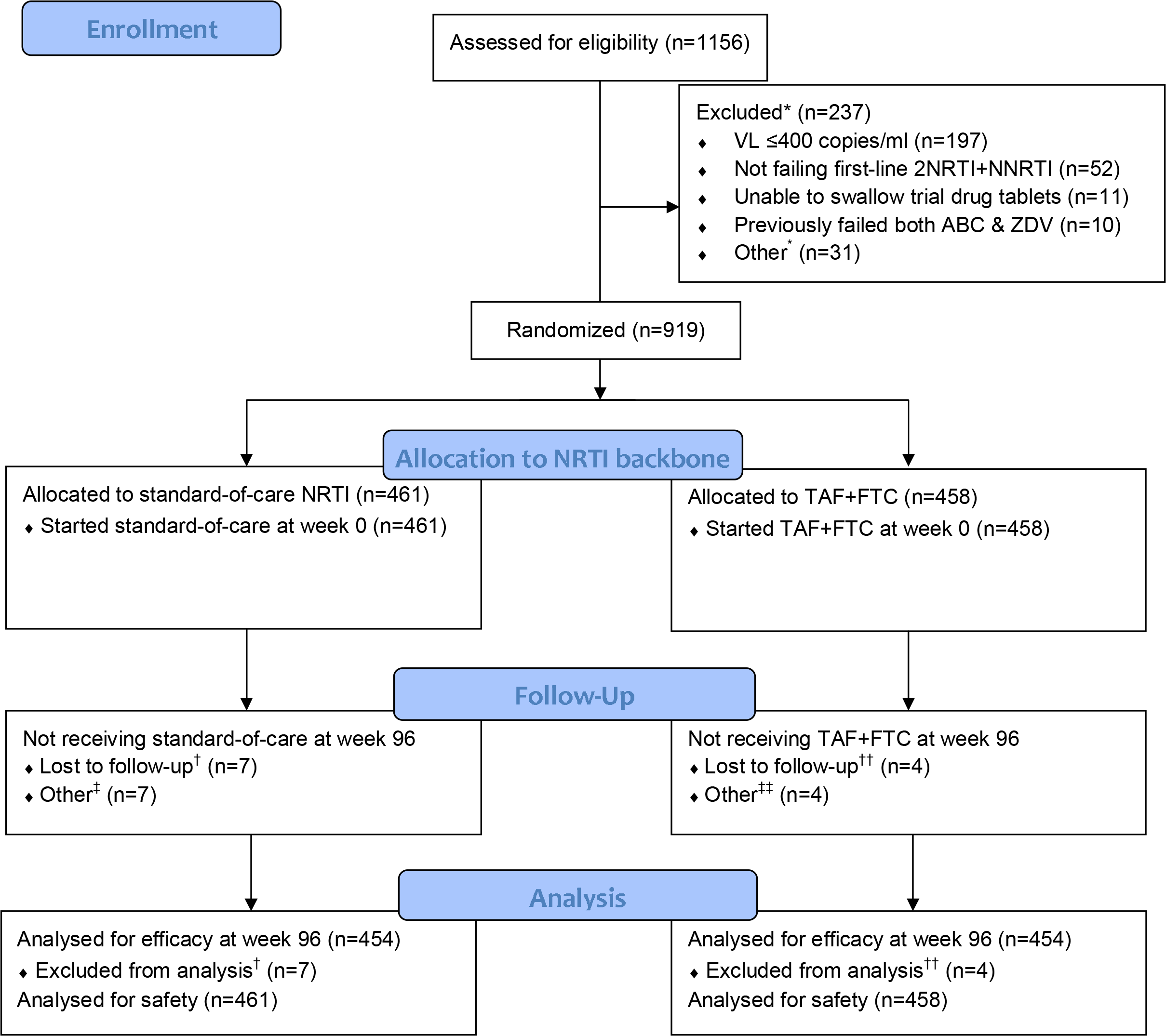
CONSORT flow diagram ABC denotes abacavir, FTC emtricitabine, NNRTI non-nucleoside reverse transcriptase inhibitor, NRTI nucleoside/nucleotide reverse transcriptase inhibitor, TAF tenofovir alafenamide fumarate, VL HIV viral load, and ZDV zidovudine. Note: All children also allocated to anchor drug ^*^ Reasons are not mutually exclusive therefore total to more than the total number of non-randomisations Other reasons: declined to participate (n=7), did not return for enrolment within window (n=4), not aged 3-15 (n=4), biochemical (n=3), previously failed ritonavir-boosted lopinavir (n=2), contraception (n=1), contraindications (n=1), co-morbidities (n=1), died (n=1), other (n=9) ^†^ Moved (n=4), social problems (n=3) ^††^ Moved (n=4) ^‡^ Started third-line (n=3), hypersensitivity (n=2), anaemia (n=1), patient decision (n=1) ^‡‡^ Patient decision (n=3), interaction (n=1)

In SOC, 217/461 (47.1%) initiated ABC/3TC and 244 (52.9%) ZDV/3TC. Over 96 weeks, 98.9% of visits were attended and only 11 children (1.2%) were lost to follow-up (Figure 1). 674 (73%) entered extended follow-up (median 60 (IQR 30,75) additional weeks). Prior to week 96, children spent 99.1% of time on allocated NRTI backbone (99.5% TAF/FTC vs. 98.8% SOC) and only five (0.5%) initiated third-line ART (2 (0.4%) TAF/FTC vs. 3 (0.7%) SOC). In extended follow-up, children spent 93.5% of time on allocated NRTI (95.6% TAF/FTC, 91.4% SOC) (Figure S1 in Supplementary Appendix 1).

Viral suppression was high in both arms (Figure 2). At week 96, 406/454 (89.4%) TAF/FTC vs. 378/454 (83.3%) SOC had VL <400copies/mL (adjusted difference +6.3% [95% confidence interval (CI) +2.0%,+10.6%]; p=0.004). Therefore, TAF/FTC was non-inferior to SOC according to the pre-specified (-)10% margin, and in fact superior. There was no evidence of heterogeneity of the effect of TAF/FTC vs. SOC in any of 11 sub-groups (p_interaction_>0.1) (Figure S2 in Supplementary Appendix 1), including first-line NRTI (ABC vs. ZDV) (p_interaction_=0.97), randomised anchor drug, country and baseline VL. Results of per-protocol analyses were similar: 403/449 (89.8%) TAF/FTC vs. 370/445 (83.1%) SOC had VL <400copies/mL (adjusted difference 6.8% [2.4%,11.1%]; p=0.002). Suppression <60 and <1000c/ml VL thresholds were similar, as were results at weeks 48 and 144 (Table S3).

**Figure 2:**
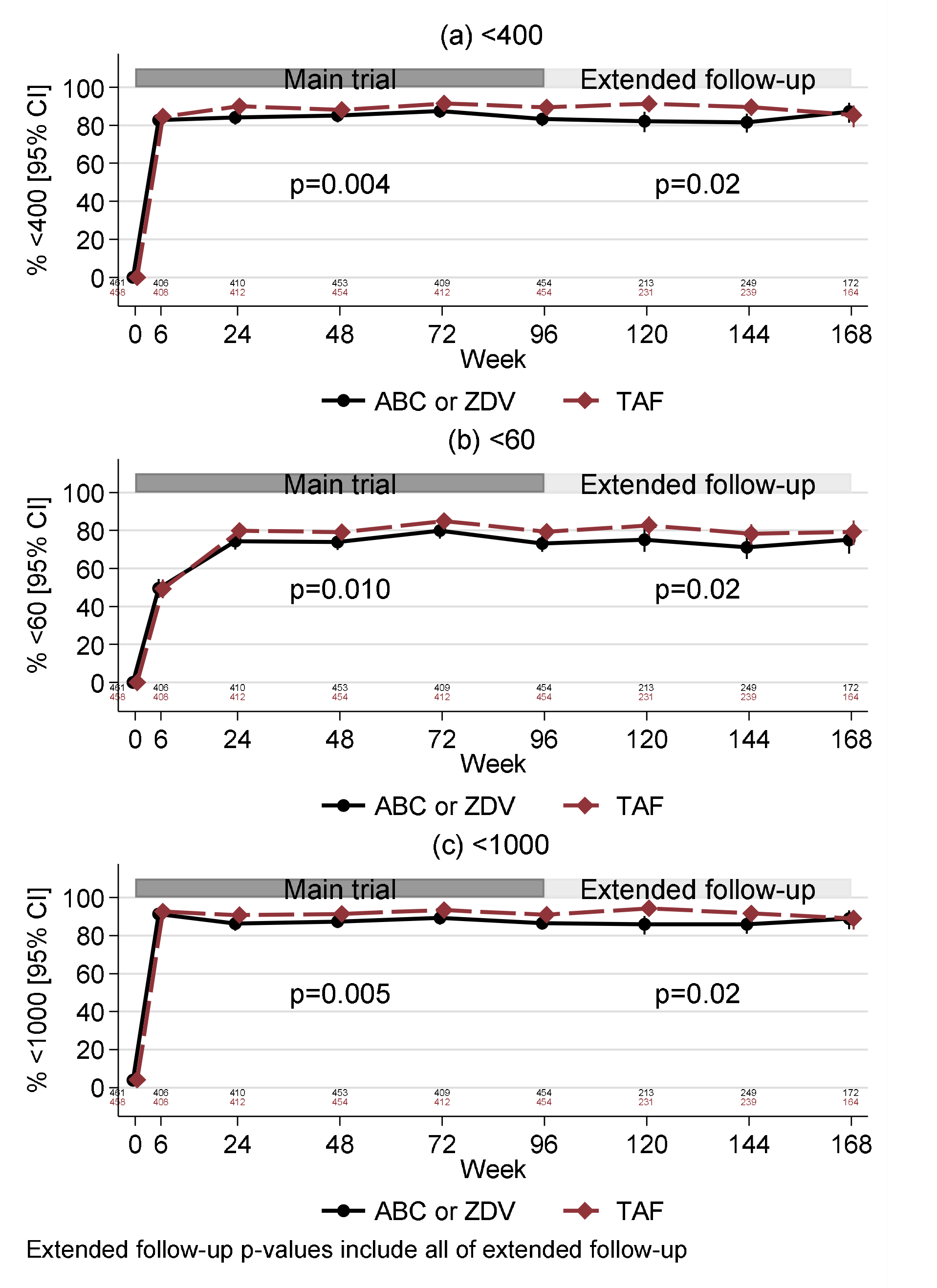
Percentage of children with HIV viral load <400 copies/ml (a), <60 copies/ml (b) and <1000 copies/ml (c), over time during the main trial and during extended follow-up ABC denotes abacavir, TAF tenofovir alafenamide fumarate and ZDV zidovudine

Over 96 weeks, there were only nine WHO stage 3/4 events and one child died (TAF/FTC, from hypotension/toxic shock secondary to severe malnutrition, judged unrelated to ART) (WHO 3/4 event/death: 5 TAF/FTC vs. 5 SOC). CD4 count improved in both arms (+103 vs. +67 cells/mm^3^ at week 96; mean difference between arms (averaged over all visits to week 96) +24 [95% CI -9,+58]), as did CD4% (+7.3% vs. +7.5%) (+0.4% [-0.4%,+1.1%]). In extended follow-up, there was no evidence of difference between arms in either CD4 or CD4% (Figure S3 in Supplementary Appendix 1).

Over 96 weeks, weight-, height- and BMI-for-age increased significantly more with TAF/FTC vs. SOC (mean difference in Z-scores between arms was +0.09 [95% CI +0.04,+0.13], 0.04 [+0.01,+0.07] and 0.10 [+0.04,+0.16], respectively); in extended follow-up, increases were generally maintained and similar (Figure S4 in Supplementary Appendix 1). Comparing TAF/FTC vs. SOC at week 96, the corresponding weight increase was 7.0 vs. 6.2kg; height increase was 10.2 vs. 9.8cm.

Over 96 weeks, 127/919 (13.8%) children experienced a total of 176 grade 3/4 adverse events (AEs) (63 (13.8%) TAF/FTC vs. 64 (13.9%) SOC) (p=0.93 Cox model) (Table 2; Table S4), including eight “specific infections”, all in SOC (4 malaria, 3 tuberculosis, 1 herpes zoster). 29 (3.2%) children experienced a total of 31 serious adverse events (SAEs) (15 (3.3%) TAF/FTC vs. 14 (3.0%) SOC) (p=0.84) (Table S5), most were hospitalisations, with intercurrent infections. 24 (2.6%) children experienced a total of 41 ART-modifying AEs (any grade) (11 (2.4%) TAF/FTC vs. 13 (2.8%) SOC) (p=0.68), of which 33 were protocol-specified modifications due to tuberculosis (Table 2).

**Table 2:**
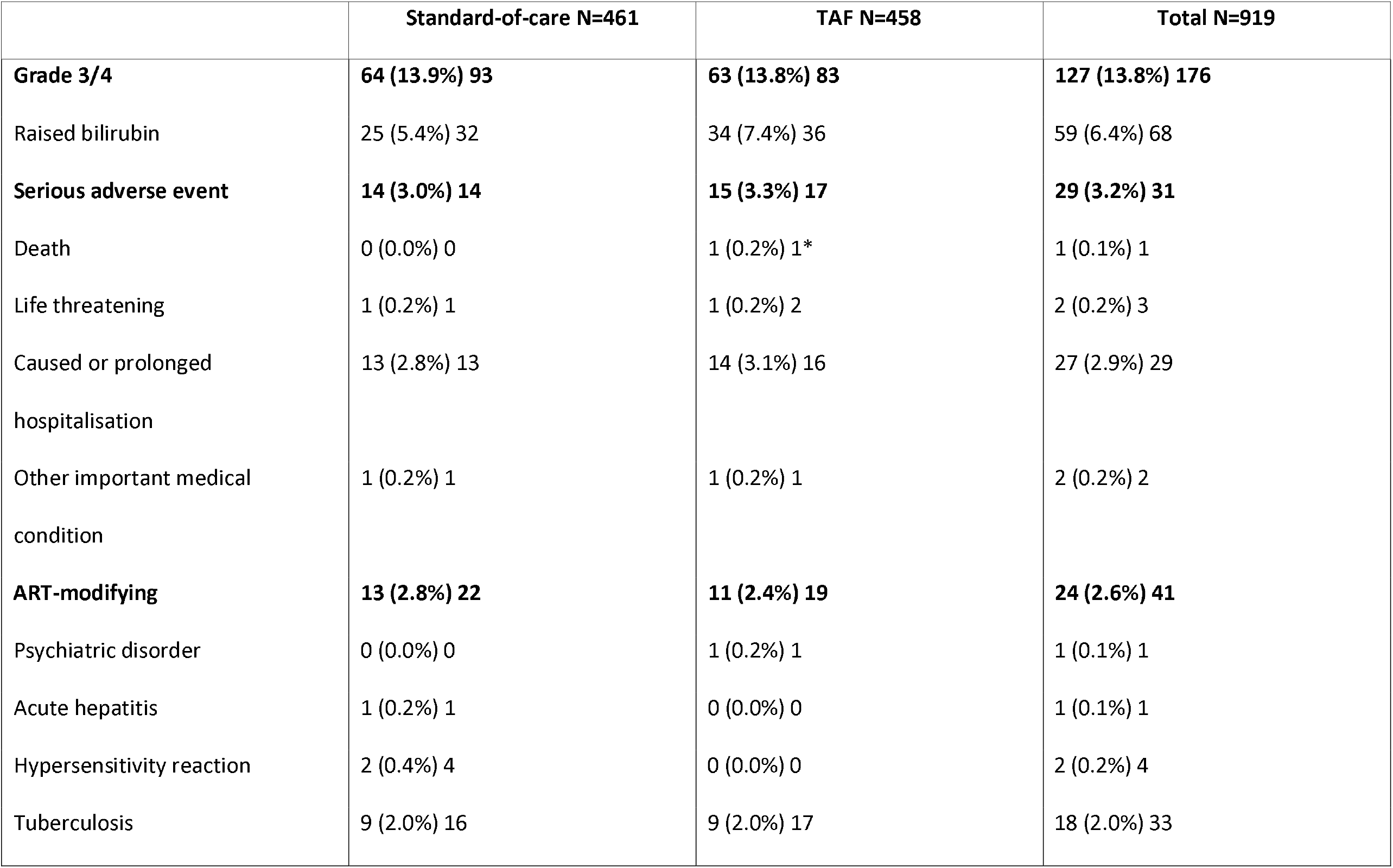
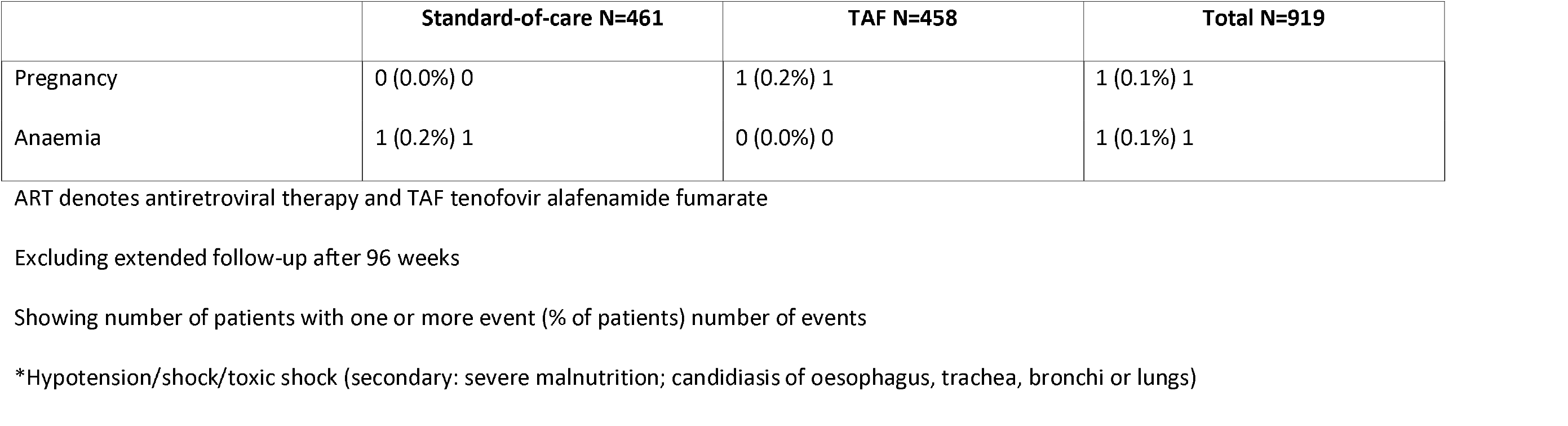
Grade 3 and 4, serious and ART-modifying adverse events during 96-week follow-up.

There was no evidence of differences in fasting lipids (total, HDL, LDL cholesterol and triglycerides) between arms over 96 weeks (p>0.1), nor in extended follow-up (p>0.05) (Figure S5 in Supplementary Appendix 1).

Calcaneal ultrasounds performed on all children showed no evidence of differences between arms over 96 weeks (p>0.3), nor in extended follow-up (p>0.2). DEXA scans performed in 170 children at weeks 0, 48 and 96 showed no evidence of differences between arms in lumbar total bone mineral content (BMC), bone mineral density (BMD) or BMD Z-score (unadjusted for height) and total body less head (TBLH) BMD Z-score (all p>0.05) TBLH BMC and BMD increased slightly more with TAF/FTC vs. SOC (p=0.02 and p=0.04, respectively; Figure 3).

**Figure 3:**
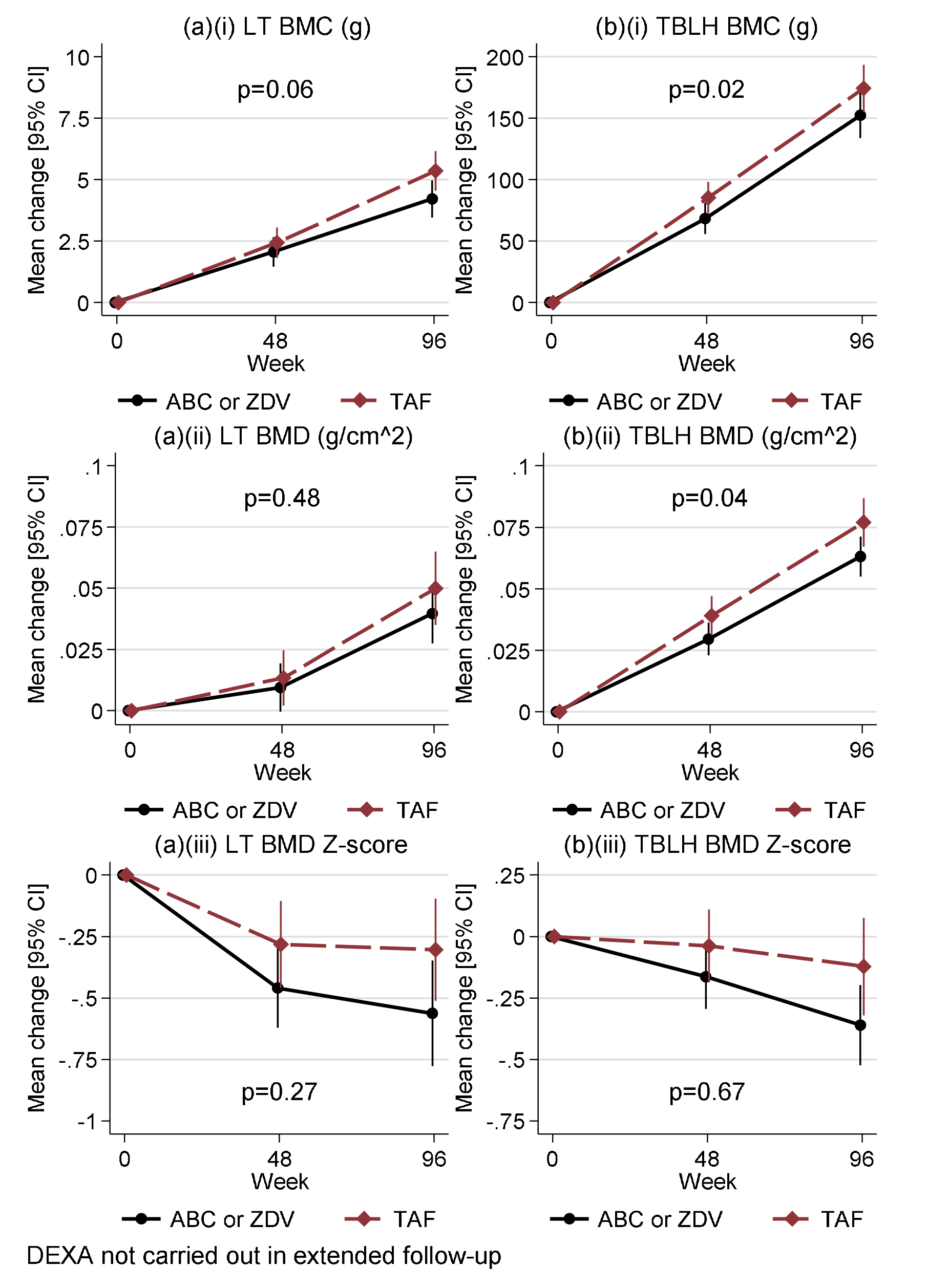
Change in (a) Lumbar total and (b) total body less head (i) bone mineral content, (ii) bone mineral density and (iii) bone mineral density Z-score ABC denotes abacavir, BMC bone mineral content, BMD bone mineral density, DEXA dual energy x-ray absorptiometry, LT lumbar total, TAF tenofovir alafenamide fumarate, TBLH total body less head and ZDV zidovudine

There was a small reduction in mean CrCl in both arms at week 96, which was greater in TAF vs. SOC (mean -16 vs. -11ml/min; p=0.0007), and which persisted in extended follow-up (p=0.001) (Figure S6 in Supplementary Appendix 1). However, no child discontinued treatment in the TAF arm for renal dysfunction. There was no evidence of differences in phosphate excretion between arms (Figure S7 in Supplementary Appendix 1).

### Cost-effectiveness Analysis

There was no significant difference in QALYs so the primary analysis focused on costs only. TAF/FTC was less costly than SOC by $37.68, with a 100% probability of being cost-saving. This saving could be used to generate 0.0754 QALY elsewhere based on a cost-effectiveness threshold of $500 per QALY. Further detail is included in Cost Effectiveness Supplementary File.

## Discussion

Although under 10% of the 1.5 million CLHIV currently on ART globally are on second-line ART,^4,5^ this proportion is likely to increase with greater access to first-line ART and VL testing. CHAPAS-4 is the first large randomized trial to evaluate NRTI backbone options including TAF for second-line ART in African children. It found that TAF/FTC provides superior viral suppression compared to current SOC of ABC/3TC or ZDV/3TC. It is also the first time a small paediatric FDC of FTC/TAF (120/15mg) has been used in children. Children did very well clinically with only one death over 96 weeks in a child with advanced disease, and very few required hospitalisation or experienced HIV disease progression. This is in part attributable to the relatively high CD4 counts at baseline and supports the principle of not delaying switch to second-line until evidence of significant immunocompromise.

The superior viral suppression of 89.4% at 96 weeks observed with TAF/FTC is comparable to the 93-100% reported in four small single-arm paediatric trials of TAF conducted in Africa, Asia and North America.^12^ Of note, over 85% were virologically suppressed at baseline in these studies, whereas all children in CHAPAS-4 had VL >400c/ml at baseline. Our results are also similar to the 86-92% viral suppression rates observed on TDF and TAF in the adult African NADIA and VISEND trials respectively,^6,7^ and the 84-86% VL suppression at 96 weeks observed in a pooled analysis of TDF/TAF in 14 adult trials.^13^

Weight-, height- and BMI-for-age z-scores increased more with TAF/FTC, suggesting overall better growth. We did not observe excessive weight gain over 96 weeks on TAF/FTC, including among children also taking DTG. The higher weight-gain observed with TAF/FTC was small in absolute terms and paralleled by an increase in height, and could be a consequence of improved virological suppression. Of note, children generally had normal or low weight and BMI-for-age at baseline, with only 5 children having a BMI for age z-score of ≥2. Among adults from the Zambian VISEND trial, greater weight gain (by∼2kg) was observed among participants (especially women) on TAF with DTG compared with ZDV with bPI or TDF with DTG.^6^

TAF was safe with no evidence of bone toxicity, and if anything, greater increases in BMD as assessed by TBLH Dexa scans vs. SOC; this was irrespective of anchor drug (DTG or bPIs). These findings, alongside the additional benefits of small pill size, once-daily administration, low cost and low risk of hypersensitivity, make TAF a valuable second-line option. Although we observed that mean CrCl decreased slightly more over 96 weeks with TAF/FTC, values remained within normal limits, with no associated grade 3/4 adverse events, and are unlikely to be clinically significant; no child discontinued medication for renal dysfunction, and phosphate excretion was not increased.

The within trial economic analysis showed that that TAF/FTC would be cost-effective compared to SOC with large cost-savings which could generate health benefits elsewhere. Development and provision of generic TAF FDC’s would potentially further enhance these benefits.

The CHAPAS-4 trial was conducted at six centres in three African countries, including three centres outside capital cities, increasing generalizability of results across sub-Saharan Africa. However, one limitation is that the trial enrolled children with NNRTI-based first-line ART failure, a situation which is becoming less common with first-line DTG rollout. Caution maybe needed when extrapolating to using TAF for first-line treatment or in children with DTG-based first-line ART failure, although it is likely that TAF would also have excellent safety and efficacy in these circumstances.^8^

In conclusion, TAF/FTC is a highly efficacious, safe and cost-effective addition to the NRTI backbone options for second-line ART for children living with HIV. Results support development of child friendly FDCs of TAF/FTC with or without anchor drugs (e.g. DTG or bPI). The results support the inclusion of TAF containing regimens on the priority list of the WHO Paediatric Drug Optimization (PADO) program,^14^ so paediatric formulations are prioritised for development, allowing them to be considered for inclusion in future WHO HIV paediatric treatment guidelines.

## Data Availability

MRC CTU at UCL supports a controlled access approach based on completion of a data request proforma available from the corresponding author (mrcctu.ucl.ac.uk/our-research/other-research-policy/data-sharing).

## Acknowledgements

We thank the participants and their families for taking part in the trial. We also acknowledge the following individuals in the partner institutions, funding bodies and pharmaceutical companies.

Clinical Trials Unit:

MRC CTU at UCL

Di Gibb, Sarah Walker, Anna Turkova, Clare Shakeshaft, Moira Spyer, Margaret Thomason, Anna Griffiths, Lara Monkiewicz, Sue Massingham, Alex Szubert, Alasdair Bamford, Katja Doerholt, Amanda Bigault, Nimisha Dudakia, Annabelle South, Nadine Van Looy, Carly Au, Hannah Sweeney

Trial Sites:

Joint Clinical Research Centre Lubowa, Uganda: Cissy M. Kityo, Victor Musiime, Eva Natukunda, Esether Nambi, Diana Rutebarika Antonia, Rashida Nazzinda, Imelda Namyalo, Joan Nangiya, Lilian Nabeeta, Aidah Nakalyango, Lilian Kobusingye, Caroline Otike, Winnie Namala, Phionah Ampaire, Ayesiga Edgar, Claire Nasaazi, Milly Ndigendawani, Paul Ociti, Priscilla Kyobutungi, Ritah Mbabazi, Phyllis Mwesigwa Rubondo, Juliet Ankunda, Mariam Naabalamba, Mary Nannungi, Alex Musiime, Faith Mbasani, Babu Enoch Louis, Josephine Namusanje, Denis Odoch, Edward Bagirigomwa, Eddie Rubanga, Disan Mulima, Paul Oronon, Eram David Williams, David Baliruno, Josephine Kobusingye, Agnes Uyungrwoth, Barbara Mukanza, Jimmy Okello, Emily Ninsiima, Lutaro Ezra, Christine Nambi, Nansaigi Mangadalen, Musumba Sharif, Nobert B. Serunjogi, Otim Thomas

Joint Clinical Research Centre Mbarara, Uganda: Abbas Lugemwa, Shafic Makumbi, Sharif Musumba, Edward Mawejje, Ibrahim Yawe, Linda Jovia Kyomuhendo, Mariam Kasozi, Rogers Ankunda, Samson kariisa, Christine Inyakuwa, Emily Ninsiima, Lorna Atwine, Beatrice Tumusiime, John Ahuura, Deogracious Tukwasibwe, Violet Nagasha, Judith Kukundakwe, Mariam Zahara Nakisekka, Ritah Winnie Nambejja, Mercy Tukamushaba, Rubinga Baker, Edridah Keminyeto, Barbara Ainebyoona, Sula Myalo, Juliet Acen, Nicholas Jinta Wangwe, Ian Natuhurira, Gershom Kananura Natukunatsa

University Teaching Hospital, Zambia: Veronica Mulenga, Chishala Chabala, Joyce Chipili Lungu, Monica Kapasa, Khozya Zyambo, Kevin Zimba, Chungu Chalilwe, Dorothy Zangata, Ellen Shingalili, Naomi Mumba, Nayunda Kaonga, Mukumbi Kabesha, Oliver Mwenechanya, Terrence Chipoya, Friday Manakalanga, Stephen Malama, Daniel Chola

Arthur Davison Children’s Hospital, Zambia: Bwendo Nduna, Mwate Mwamabazi, Kabwe Banda, Beatrice Kabamba, Muleya Inambao, Pauline Mahy Mukandila, Mwizukanji Nachamba, Stella Himabala, Shadrick Ngosa, Davies Sondashi, Collins Banda, Mark Munyangabe, Grace Mbewe Ngoma, Sarah Chimfwembe, Mercy Lukonde Malasha, Mumba Kajimalwendo, Henry Musukwa, Shadrick Mumba

University of Zimbabwe Clinical Research Centre, Zimbabwe: James Hakim, Mutsa Bwakura-Dangarembizi, Kusum Nathoo, Taneal Kamuzungu, Ennie Chidziva, Joyline Bhiri, Joshua Choga, Hilda Angela Mujuru, Godfrey Musoro, Vivian Mumbiro, Moses Chitsamatanga, Constantine Mutata, Shepherd Mudzingwa, Secrecy Gondo, Columbus Moyo, Ruth Nhema, Kathryn Boyd, Farai Matimba, Vinie Kouamou, Richard Matarise, Zorodzai Tangwena, Taona Mudzviti, Allen Matubu, Alfred Kateta, Victor Chinembiri, Dorinda Mukura, Joy Chimanzi, Dorothy Murungu, Wendy Mapfumo, Pia Ngwaru, Lynette Chivere, Prosper Dube, Trust Mukanganiki, Sibusisiwe Weza, Tsitsi Gwenzi, Shirley Mutsai, Misheck Phiri, Makhosonke Ndlovu, Tapiwa Gwaze, Stuart Chitongo, Winisayi Njaravani, Sandra Musarurwa, Cleopatra Langa, Sue Tafeni, Wilbert Ishemunyoro, Nathalie Mudzimirema

Mpilo Central Hospital, Zimbabwe: Wedu Ndebele, Mary Nyathi, Grace Siziba, Getrude Tawodzera, Tracey Makuchete, Takudzwa Chidarura, Shingaidzo Murangandi, Lawrence Mafaro, Owen Chivima, Sifiso Dumani, Beaullar Mampondo, Constance Maphosa, Debra Mwale, Rangarirai Dhlamini, Thabani Sibanda, Nobukhosi Madubeko, Silibaziso Nyathi, Zibusiso Matiwaza, Blessing Sanyanga, Prince Ziyera, Gamuchirai Mauro, Titshabona Ncube, Again Gwapedza, Davison Mashoko

Local External Site Monitors

Uganda: Sylvia Nabukenya, Harriet Tibakabikoba, Sarah Nakalanzi, Cynthia Williams

Zimbabwe: Precious Chandiwana, Winnie Gozhora, Benedictor Dube

Zambia: Sylvia Mulambo, Hope Mwanyungwi

Sub-studies

PK sub-studies – Radboud University Medical Centre: David Burger, Angela Colbers, Hylke Waalewijn, Lisanne Bevers, Shaghayegh Mohsenian-Naghani, Anne Kamphuis

PK sub-studies – University of Cape Town: Helen McIlleron, Jennifer Norman, Lubbe Wiesner, Roeland Wasmann, Paolo Denti, Lufina Tsirizani Galileya

Toxicity sub-study: Eva Natukunda, Victor Musiime, Phillipa Musoke

Health Economics sub-study – University of York: Paul Revill, Simon Walker, Yingying Zhang

Trial Committees

Independent Trial Steering Committee Members: Adeodata Kekitiinwa, Angela Mushavi, Febby Banda Kawamya, Denis Tindyebwa, Hermione Lyall, Ian Weller

Independent Data Monitoring Committee Members: Tim Peto, Philippa Musoke, Margaret Siwale, Rose Kambarami

## Funders

EDCTP: Johanna Roth, Pauline Beattie

Janssen Pharmaceuticals

Gilead Sciences

Drug Contributors

Gilead Sciences Ltd., Janssen Pharmaceuticals, Viiv Healthcare, GSK Ltd., Cipla Ltd.

## Ethical approval

The trial was approved by ethics committees in Uganda (Joint Research Ethics Committee (JREC)), Zambia (University of Zambia Biomedical Research Ethics Committee (UNZABREC)), Zimbabwe (Joint Research Ethics Committee University of Zimbabwe College of Health Sciences (JREC), Research Council Zimbabwe (RCZ)), South Africa (University of Cape Town Human Research Ethics Committee) and United Kingdom (UCL Research Ethics Committee).

