## Supplemental File 1 for "Second-line tenofovir alafenamide for children with HIV in Africa"

**Table S1: Weight-band based dosing of trial NRTI backbones.**

| | ABC/3TC | | ZDV/3TC | | FTC/TAF | |
|---|---|---|---|---|---|---|
| | 120/60 mg | 600/300 mg | 60/30 mg | 300/150 mg | 120/15 mg | 200/25 mg |
| **WHO weight bands** | OD | OD | BD (am+pm) | BD (am+pm) | OD | OD |
| **14-19.9 kg** | 2.5 | - | 3+2* | - | 1 | - |
| **20-24.9 kg** | 3 | - | 3+3 | - | 1 | - |
| **25-34.9 kg** | - | 1 | - | 1+1 | - | 1 |
| **35- kg (adult)** | - | 1 | - | 1+1 | - | 1 |

ABC denotes abacavir, BD twice daily, FTC emtricitabine, NRTI nucleoside/nucleotide reverse transcriptase inhibitor, OD once daily, TAF tenofovir alafenamide fumarate, ZDV zidovudine and 3TC lamivudine

**Table S2: Additional baseline characteristics**

| | Standard-of-care N=461 | TAF N=458 | Total N=919 |
|---|---|---|---|
| Centre | | | |
| Uganda/Kampala | 100 (21.7%) | 101 (22.1%) | 201 (21.9%) |
| Uganda/Mbarara | 99 (21.5%) | 97 (21.2%) | 196 (21.3%) |
| Zambia/Lusaka | 60 (13.0%) | 61 (13.3%) | 121 (13.2%) |
| Zambia/Ndola | 37 (8.0%) | 37 (8.1%) | 74 (8.1%) |
| Zimbabwe/Harare | 109 (23.6%) | 110 (24.0%) | 219 (23.8%) |
| Zimbabwe/Bulawayo | 56 (12.1%) | 52 (11.4%) | 108 (11.8%) |

Values are n (%) or median (IQR). TAF denotes tenofovir alafenamide fumarate

**Table S3: Viral load comparisons at weeks 48, 96 and 144 for thresholds of viral load <400, <60 and <1000 copies/ml**

| Comparison | Week 48 (n=907): difference (%) [95% CI] | p | Week 96 (n=908): difference (%) [95% CI] | p | Week 144 (n=488): difference (%) [95% CI] | p |
|---|---|---|---|---|---|---|
| **<400 copies/ml** | | | | | | |
| TAF vs. Standard-of-care | 3.0 [-1.3, 7.4] | 0.17 | 6.3 [2.0, 10.6] | 0.004 | 8.2 [2.1, 14.3] | 0.008 |
| **<60 copies/ml** | | | | | | |
| TAF vs. Standard-of-care | 5.3 [0.0, 10.7] | 0.0499 | 6.3 [1.0, 11.5] | 0.02 | 7.2 [-0.2, 14.5] | 0.06 |
| **<1000 copies/ml** | | | | | | |
| TAF vs. Standard-of-care | 4.1 [0.1, 8.0] | 0.04 | 4.6 [0.6, 8.6] | 0.02 | 5.8 [0.4, 11.3] | 0.04 |

CI denotes confidence interval and TAF tenofovir alafenamide fumarate

**Table S4: Grade 3 and 4 adverse events during 96-week follow-up**

| | Standard-of-care N=461 | TAF N=458 | Total N=919 | p* |
|---|---|---|---|---|
| **Any** | **64 (13.9%) 93** | **63 (13.8%) 83** | **127 (13.8%) 176** | **1.00** |
| **CNS** | **0 (0.0%) 0** | **1 (0.2%) 3** | **1 (0.1%) 3** | **0.50** |
| **Psychiatric** | **3 (0.7%) 3** | **2 (0.4%) 2** | **5 (0.5%) 5** | **1.00** |
| **Lower Respiratory Tract** | **2 (0.4%) 2** | **3 (0.7%) 4** | **5 (0.5%) 6** | **0.69** |
| **Cardiovascular** | **0 (0.0%) 0** | **1 (0.2%) 1** | **1 (0.1%) 1** | **0.50** |
| **Eye** | **1 (0.2%) 1** | **0 (0.0%) 0** | **1 (0.1%) 1** | **1.00** |
| **Gastrointestinal** | **0 (0.0%) 0** | **2 (0.4%) 2** | **2 (0.2%) 2** | **0.25** |
| **Hepatic** | **4 (0.9%) 4** | **2 (0.4%) 2** | **6 (0.7%) 6** | **0.69** |
| **Musculoskeletal** | **0 (0.0%) 0** | **1 (0.2%) 1** | **1 (0.1%) 1** | **0.50** |
| **Skin** | **4 (0.9%) 5** | **1 (0.2%) 2** | **5 (0.5%) 7** | **0.37** |
| **Haematological** | **25 (5.4%) 31** | **18 (3.9%) 22** | **43 (4.7%) 53** | **0.35** |
| Pancytopenia, bone marrow depression | 0 (0.0%) 0 | 1 (0.2%) 2 | 1 (0.1%) 2 | |
| Anaemia with clinical symptoms | 6 (1.3%) 6 | 1 (0.2%) 1 | 7 (0.8%) 7 | |
| Thrombocytopenia | 3 (0.7%) 3 | 4 (0.9%) 6 | 7 (0.8%) 9 | |
| Leucopenia | 1 (0.2%) 1 | 0 (0.0%) 0 | 1 (0.1%) 1 | |
| Neutropenia | 10 (2.2%) 10 | 5 (1.1%) 6 | 15 (1.6%) 16 | |
| Anaemia with no clinical symptoms | 8 (1.7%) 8 | 5 (1.1%) 5 | 13 (1.4%) 13 | |
| Lymphopenia | 3 (0.7%) 3 | 2 (0.4%) 2 | 5 (0.5%) 5 | |
| **Biochemical** | **28 (6.1%) 35** | **36 (7.9%) 39** | **64 (7.0%) 74** | **0.30** |
| Raised liver enzymes | 0 (0.0%) 0 | 1 (0.2%) 1 | 1 (0.1%) 1 | |
| Raised AST | 2 (0.4%) 2 | 1 (0.2%) 1 | 3 (0.3%) 3 | |
| Raised ALT | 1 (0.2%) 1 | 1 (0.2%) 1 | 2 (0.2%) 2 | |
| Raised bilirubin | 25 (5.4%) 32 | 34 (7.4%) 36 | 59 (6.4%) 68 | |
| **Systemic** | **1 (0.2%) 1** | **2 (0.4%) 2** | **3 (0.3%) 3** | **0.62** |
| **Specific Infections** | **8 (1.7%) 8** | **0 (0.0%) 0** | **8 (0.9%) 8** | **0.008** |
| Herpes Zoster (Varicella Zoster) – cutaneous | 1 (0.2%) 1 | 0 (0.0%) 0 | 1 (0.1%) 1 | |
| Tuberculosis – disseminated/miliary | 1 (0.2%) 1 | 0 (0.0%) 0 | 1 (0.1%) 1 | |
| P. falciparum malaria | 4 (0.9%) 4 | 0 (0.0%) 0 | 4 (0.4%) 4 | |
| Tuberculosis – abdominal | 2 (0.4%) 2 | 0 (0.0%) 0 | 2 (0.2%) 2 | |
| **Undiagnosed Fevers** | **0 (0.0%) 0** | **1 (0.2%) 1** | **1 (0.1%) 1** | **0.50** |
| **Tumours** | **1 (0.2%) 1** | **0 (0.0%) 0** | **1 (0.1%) 1** | **1.00** |

| | Standard-of-care N=461 | TAF N=458 | Total N=919 | p* |
|---|---|---|---|---|
| **Pregnancy Associated** | **0 (0.0%) 0** | **1 (0.2%) 1** | **1 (0.1%) 1** | **0.50** |
| **Other** | **2 (0.4%) 2** | **1 (0.2%) 1** | **3 (0.3%) 3** | **1.00** |

Excluding extended follow-up after 96 weeks
Detail within body system provided where p≤0.05 or ≥10% of children experienced an event
Showing number of patients with one or more event (% of patients) number of events
TAF denotes tenofovir alafenamide fumarate
*Fisher's exact test

**Table S5: Serious Adverse Events (SAEs) during 96-week follow-up**

| | Standard-of-care N=461 | TAF N=458 | Total N=919 | p* |
|---|---|---|---|---|
| **Any** | **14 (3.0%) 14** | **15 (3.3%) 17** | **29 (3.2%) 31** | **0.85** |
| CNS | 0 (0.0%) 0 | 1 (0.2%) 2 | 1 (0.1%) 2 | 0.50 |
| Upper Respiratory Tract | 0 (0.0%) 0 | 2 (0.4%) 2 | 2 (0.2%) 2 | 0.25 |
| Lower Respiratory Tract | 2 (0.4%) 2 | 4 (0.9%) 4 | 6 (0.7%) 6 | 0.45 |
| Gastrointestinal | 0 (0.0%) 0 | 2 (0.4%) 2 | 2 (0.2%) 2 | 0.25 |
| Hepatic | 0 (0.0%) 0 | 1 (0.2%) 1 | 1 (0.1%) 1 | 0.50 |
| Skin | 3 (0.7%) 3 | 0 (0.0%) 0 | 3 (0.3%) 3 | 0.25 |
| Haematological | 2 (0.4%) 2 | 2 (0.4%) 3 | 4 (0.4%) 5 | 1.00 |
| Systemic | 1 (0.2%) 1 | 0 (0.0%) 0 | 1 (0.1%) 1 | 1.00 |
| Specific Infections | 5 (1.1%) 5 | 2 (0.4%) 2 | 7 (0.8%) 7 | 0.45 |
| Undiagnosed Fevers | 0 (0.0%) 0 | 1 (0.2%) 1 | 1 (0.1%) 1 | 0.50 |
| Other | 1 (0.2%) 1 | 0 (0.0%) 0 | 1 (0.1%) 1 | 1.00 |

Excluding extended follow-up after 96 weeks
Detail within body system provided where p≤0.05 or ≥10% of children experienced an event
Number of patients with one or more episode (% of patients) number of episodes
TAF denotes tenofovir alafenamide fumarate
*Fisher's exact test

**Figure S1: Treatment received over time from randomization (extended follow-up from week 120-168)**

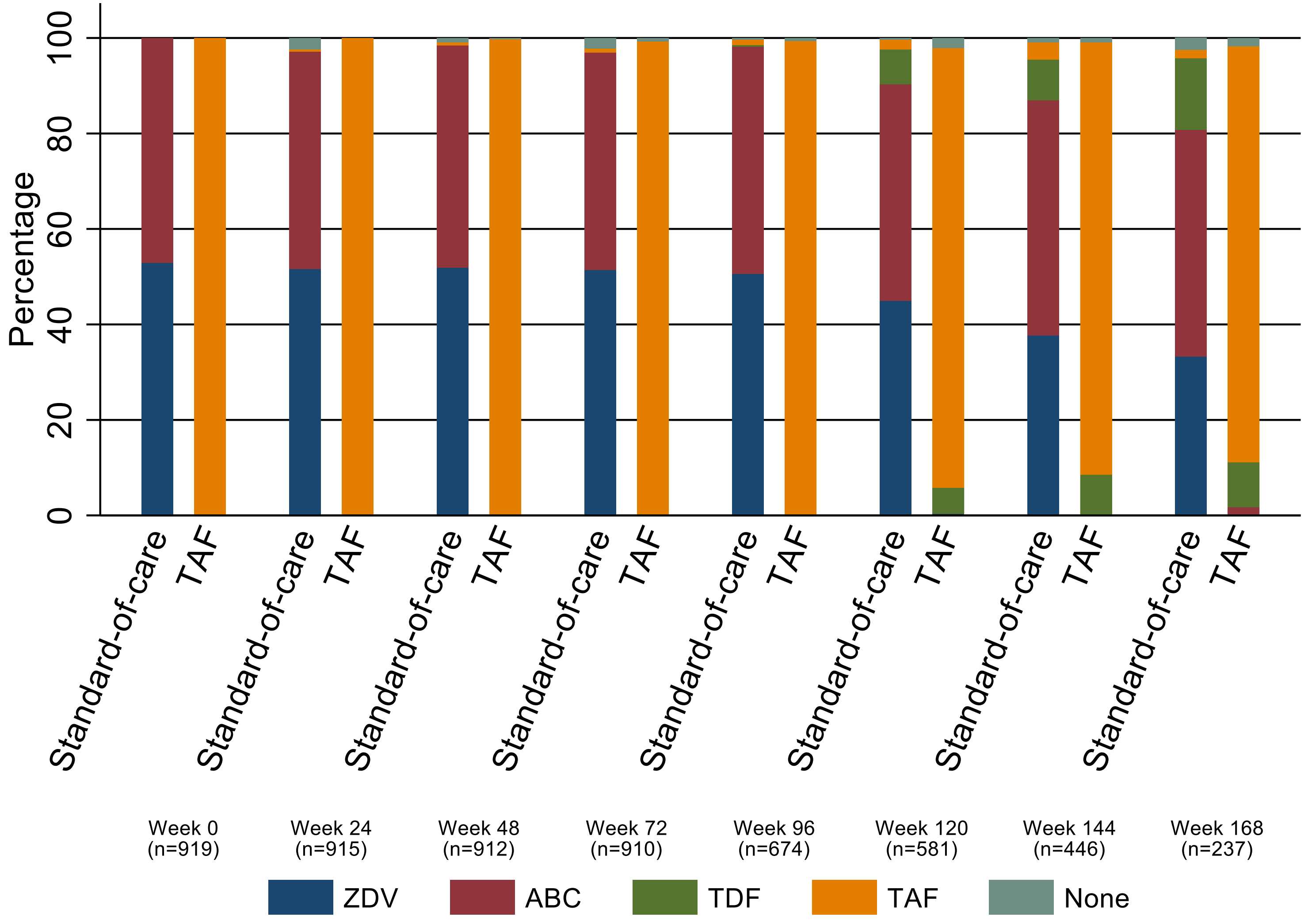

ABC denotes abacavir, TAF tenofovir alafenamide fumarate, TDF tenofovir disoproxil fumarate and ZDV zidovudine

### Figure S2: Subgroup analyses for the primary endpoint, viral load suppression <400 copies/ml at week 96

| | Standard-of-care | TAF | TAF-Standard-of-care difference (%) [95% CI] | Interaction p |
|---|---|---|---|---|
| **01 PARTICIPANT HAS BEEN RANDOMISED TO: 3RD DRUG RANDOMISATION** | | | | |
| LPV/r | 83/112(74.1%) | 97/111(87.4%) | 8.2 [3.0, 13.4] | 0.61 |
| ATV/r | 91/113(80.5%) | 102/116(87.9%) | 5.6 [-0.6, 11.9] | |
| DRV/r | 98/113(86.7%) | 105/117(89.7%) | 3.1 [-4.8, 11.0] | |
| DTG | 106/116(91.4%) | 102/110(92.7%) | 2.0 [-8.0, 12.0] | |
| **02 SITE** | | | | |
| Kampala | 94/99(94.9%) | 96/100(96.0%) | 2.5 [-10.5, 15.6] | 0.53 |
| Mbarara | 79/97(81.4%) | 90/97(92.8%) | 9.1 [3.5, 14.7] | |
| Lusaka | 39/57(68.4%) | 52/61(85.2%) | 8.4 [2.7, 14.2] | |
| Ndola | 27/37(73.0%) | 28/36(77.8%) | 2.7 [-7.5, 12.9] | |
| Harare | 90/108(83.3%) | 96/109(88.1%) | 4.0 [-3.1, 11.1] | |
| Bulawayo | 49/56(87.5%) | 44/51(86.3%) | -1.2 [-14.6, 12.2] | |
| **03 FAILING FIRST-LINE NRTI** | | | | |
| ABC | 188/239(78.7%) | 211/244(86.5%) | 5.9 [1.1, 10.7] | 0.97 |
| ZDV | 190/215(88.4%) | 195/210(92.9%) | 5.5 [-0.5, 11.5] | |
| **04 VL AT TRIAL ENTRY (COPIES/ML)** | | | | |
| <10000 | 133/161(82.6%) | 143/164(87.2%) | 3.8 [-2.2, 9.9] | 0.35 |
| 10000-99999 | 190/221(86.0%) | 208/229(90.8%) | 5.1 [-0.6, 10.7] | |
| >=100000 | 55/72(76.4%) | 55/60(91.7%) | 9.7 [4.2, 15.2] | |
| **05 COUNTRY** | | | | |
| Uganda | 173/196(88.3%) | 186/197(94.4%) | 7.6 [1.9, 13.3] | 0.45 |
| Zambia | 66/94(70.2%) | 80/97(82.5%) | 6.6 [1.1, 12.2] | |
| Zimbabwe | 139/164(84.8%) | 140/160(87.5%) | 2.5 [-4.0, 9.0] | |

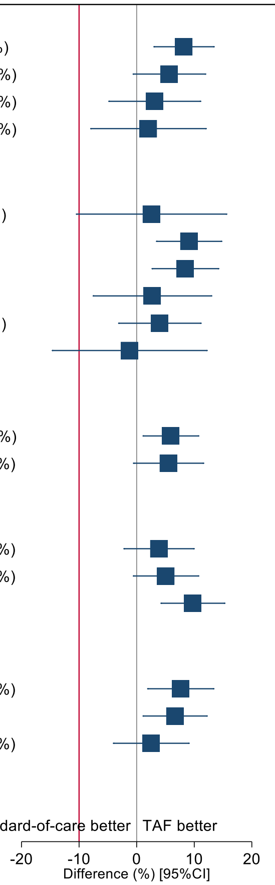

Red line denotes non-inferiority margin (10%)

| | Standard-of-care | TAF | TAF-Standard-of-care difference (%) [95% CI] | Interaction p |
|---|---|---|---|---|
| **06 AGE AT LAST BIRTHDAY (YEARS)** | | | | |
| 3-4 | 18/21(85.7%) | 16/17(94.1%) | 8.1 [-4.8, 21.0] | 0.42 |
| 5-9 | 151/172(87.8%) | 169/178(94.9%) | 8.7 [3.1, 14.2] | |
| 10-15 | 209/261(80.1%) | 221/259(85.3%) | 4.1 [-0.8, 8.9] | |
| **07 SEX** | | | | |
| Male | 205/250(82.0%) | 216/238(90.8%) | 7.8 [3.0, 12.7] | 0.21 |
| Female | 173/204(84.8%) | 190/216(88.0%) | 3.0 [-2.9, 8.9] | |
| **08 WEIGHT (KG)** | | | | |
| <20 | 91/104(87.5%) | 82/86(95.3%) | 8.9 [2.4, 15.4] | 0.67 |
| 20-<25 | 91/107(85.0%) | 105/114(92.1%) | 6.8 [0.2, 13.4] | |
| 25-<35 | 125/148(84.5%) | 151/171(88.3%) | 3.5 [-2.9, 10.0] | |
| >=35 | 71/95(74.7%) | 68/83(81.9%) | 4.4 [-2.2, 11.0] | |
| **09 FAILING FIRST LINE NNRTI** | | | | |
| EFV | 191/243(78.6%) | 227/264(86.0%) | 5.6 [0.8, 10.3] | 0.61 |
| NVP | 187/211(88.6%) | 179/190(94.2%) | 7.1 [1.2, 12.9] | |
| **10 WEIGHT-FOR-AGE Z-SCORE*** | | | | |
| <=-2.1 | 119/147(81.0%) | 134/154(87.0%) | 4.8 [-1.1, 10.7] | 0.96 |
| >-2.1 to <=-1.13 | 139/168(82.7%) | 125/140(89.3%) | 5.6 [-0.3, 11.5] | |
| >-1.13 | 120/139(86.3%) | 147/160(91.9%) | 5.9 [-0.6, 12.3] | |
| **11 CD4 AT TRIAL ENTRY (CELLS/MM^3)*** | | | | |
| <=499 | 120/153(78.4%) | 121/144(84.0%) | 4.0 [-1.9, 9.8] | 0.12 |
| >499 - <=852 | 124/153(81.0%) | 140/151(92.7%) | 9.8 [4.8, 14.8] | |
| >852 | 131/145(90.3%) | 136/150(90.7%) | 0.4 [-8.5, 9.3] | |

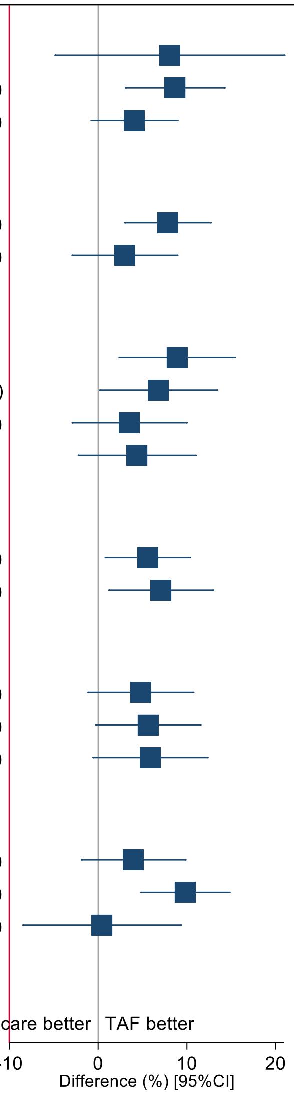

Red line denotes non-inferiority margin (10%)
*Terciles

ABC denotes abacavir, ATV/r ritonavir-boosted atazanavir, DRV/r ritonavir-boosted darunavir, DTG dolutegravir, EFV efavirenz, LPV/r ritonavir-boosted lopinavir, NNRTI non-nucleoside reverse transcriptase inhibitor, NRTI nucleoside/nucleotide reverse transcriptase inhibitor, NVP nevirapine, TAF tenofovir alafenamide fumarate, VL HIV viral load and ZDV zidovudine

**Figure S3: Change in (a) CD4 and (b) CD4% during main trial and extended follow-up**

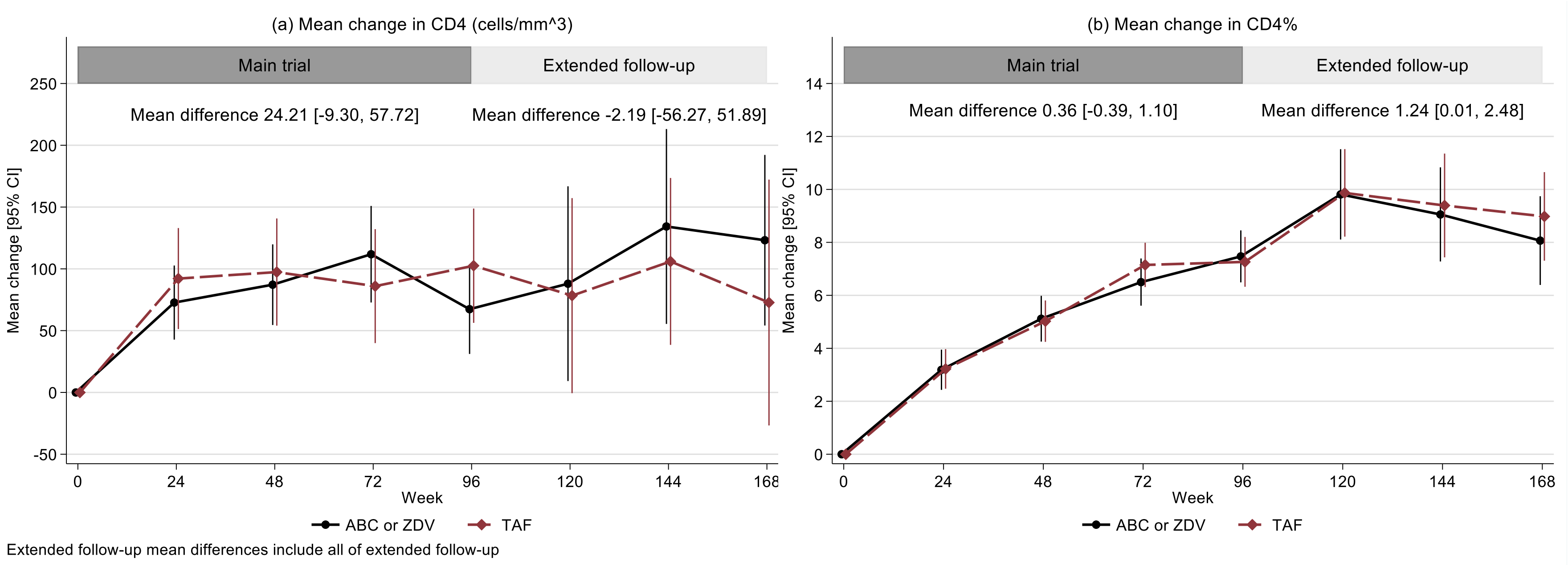

ABC denotes abacavir, TAF tenofovir alafenamide fumarate and ZDV zidovudine

**Figure S4: Change in (a) weight-, (b) height- and (c) BMI-for-age during main trial and extended follow-up**

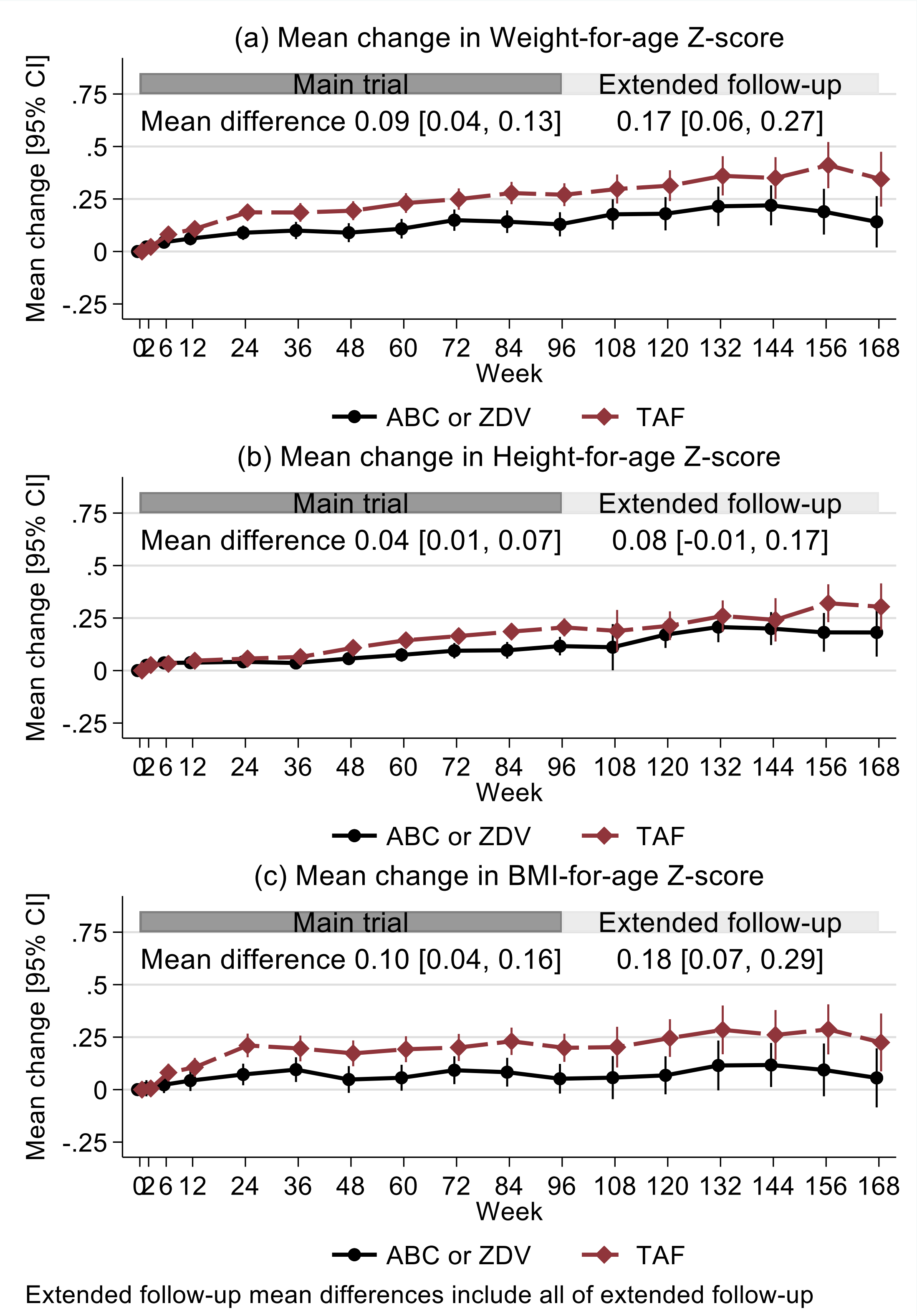

ABC denotes abacavir, BMI body mass index, TAF tenofovir alafenamide fumarate and ZDV zidovudine

**Figure S5: Change in (a) total, (b) HDL and (c) LDL cholesterol and (d) triglycerides during main trial and extended follow-up**

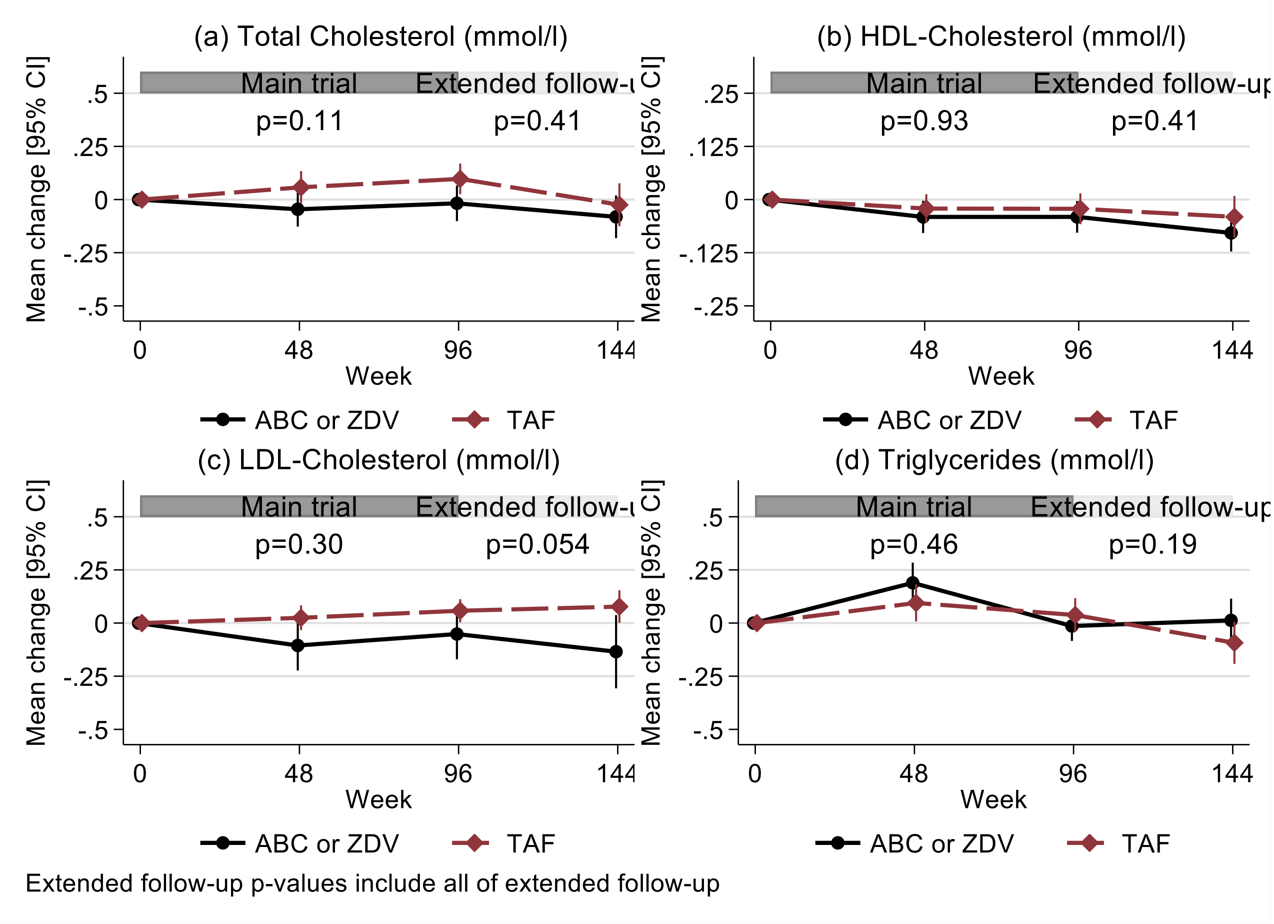

ABC denotes abacavir, HDL high-density lipoprotein, LDL low-density lipoprotein, TAF tenofovir alafenamide fumarate and ZDV zidovudine

**Figure S6: Change in creatinine clearance over main trial and extended follow up**

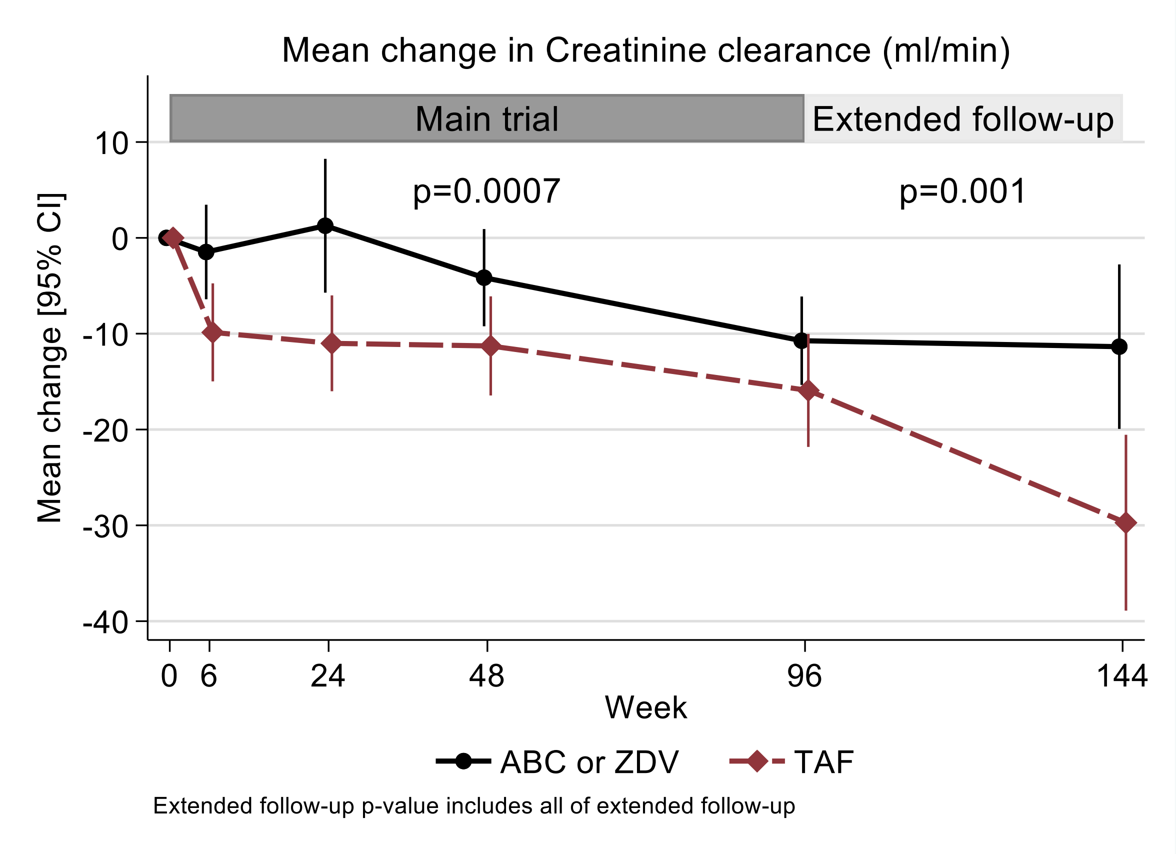

ABC denotes abacavir, TAF tenofovir alafenamide fumarate and ZDV zidovudine

**Figure S7: Change in phosphate excretion over 96 weeks**

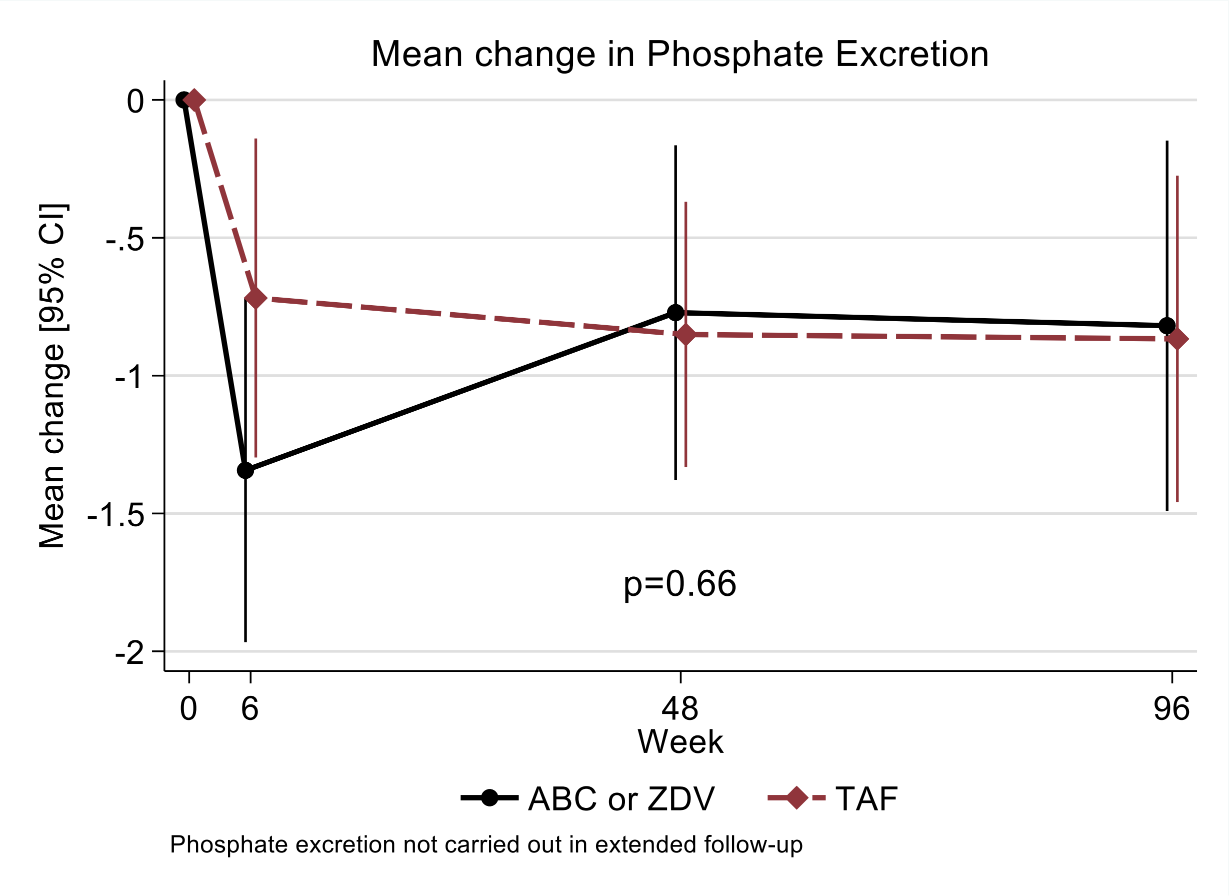

ABC denotes abacavir, TAF tenofovir alafenamide fumarate and ZDV zidovudine

**Panel S1. The effect of the COVID-19 pandemic**

At the end of March 2020, the trial sponsor and the national authorities made the decision to halt recruitment in all sites due to the COVID-19 pandemic. All enrolled participants continued to be supplied with trial medication and were followed up either at the trial clinic, at home or via phone calls, depending on the level of local lockdown. This had cost and resource implications for the sites, including additional transport and personal protective equipment (PPE). From June 2020, sites restarted recruitment following review of national guidelines and local mitigation plans. The visit window allowed was increased during periods of lockdown or travel/transport restrictions to allow safety and endpoint tests to be conducted. COVID-19 specific protocol deviations were reviewed regularly, and the impact assessed. A manual of operations as well as COVID-19 risk management plans for each site were developed to guide these processes.

Delays in recruitment that arose from this temporary pause in enrolment and the challenges posed by the implementation of 2019 WHO recommendations, which required drug optimization to dolutegravir (DTG) based regimens for all children on first and second-line ART. The independent data monitoring committee (IDMC) and trial steering committee (TSC) agreed to the proposal by trial management group (TMG) that the sample size could be reduced to 920 from 1000 whilst retaining statistical power. This was possible due to the very small loss to follow up (0.5%).
