## Supplemental File 2 for "Second-line tenofovir alafenamide for children with HIV in Africa"

**Cost-Effectiveness Analysis for Nucleos(t)ide Reverse Transcriptase Inhibitor (NRTI) Backbone Drugs in the CHAPAS-4 Trial**

Table CES6. Cost-effectiveness of SOC versus TAF/FTC ……………………………………..8

**Methods**

The health economics analysis assessed the cost-effectiveness of tenofovir alafenamide fumarate(TAF)/emtricitabine(FTC) compared to standard-of-care(SOC). Health was captured using quality-adjusted life years (QALYs) based on the EQ-5D-3L instrument and costs were captured from a health care perspective and included antiretroviral (ART) drugs, clinic visits and hospital stays. The time horizon of the analysis was restricted to the 96-week trial period and a discount rate of 3% was applied to costs and QALYs in the second year. All costs were estimated in 2022 US$. Unit costs for the analysis are reported in Tables CES1 and CES2.

The incremental cost of TAF/FTC compared to SOC was estimated by the generalized linear models with a gamma distribution and an identity link function adjusting for stratification factors, and the probabilities of SOC and TAF/FTC being least costly were estimated using probabilistic sensitivity analysis. The incremental QALY were estimated by ordinary least squares controlling for the same set of stratification factors and the baseline EQ5D indices.

**Results**

Unit costs of ART drugs and of other health care are reported in Table CES1 and CES2. Table S3 and S4 report the resources used, and costs and QALYs by randomization groups. The main health economics analysis focused on comparing the mean total costs of SOC and TAF/FTC, including costs of ART drugs, clinic visits and hospital stays, from the health system perspective as QALYs outcomes calculated based on the EQ-5D-3L instrument were not significantly different between the two nucelos(t)ide reverse transcriptase inhibitor (NRTI) backbone randomization arms with only small differences observed and no systematic trend over time. Table CES5 reports the estimated total costs of SOC and TAF/FTC, the estimated incremental cost of TAF/FTC, and the probability of each being least costly. TAF/FTC was found to be $37.68 less costly than SOC. Probabilistic sensitivity analysis suggested the probability of TAF/FTC being cost saving compared to SOC was 100%.

Table CES6 reports the full cost-effectiveness analysis including the estimated total QALYs of SOC and TAF/FTC, and incremental cost-effectiveness ratios (ICERs) and incremental net health benefits (INHBs) of TAF/FTC compared to SOC. TAF/FTC was less costly ($37.68) than SOC, it was also marginally less effective (0.0048 QALYs). One QALY loss from switching from SOC to TAF/FTC generated $7,861 of cost savings, considerably higher than the highest cost-effectiveness threshold of $500 per QALY. The INHB of TAF/FTC compared to SOC was 0.0707, using the cost-effectiveness threshold of $500 per QALY. The positive net benefit suggested that TAF/FTC was cost-effective compared to SOC. The probability of TAF/FTC being cost-effective was 100% using three different threshold levels of £100, £300 and £500 per QALY.

Table CES1: Costs of antiretroviral drugs.

| | | WHO WEIGHT BANDS | | | |
|---|---|---|---|---|---|
| | | **14-19.9 kg** | **20-24.9 kg** | **25-34.9 kg** | **35- kg (adult)** |
| **Abacavir/ Lamivudine ABC/3TC** | Daily dose | 300/150 mg | 360/180 mg | 600/300 mg | 600/300 mg |
| | Tablets taken | 2.5*120/60 mg ABC/3TC | 3*120/60 mg ABC/3TC | 1*600/300 mg[1] ABC/3TC | 1*600/300 mg[1] ABC/3TC |
| | **Unit cost($)** | **ABC/3TC (120/60 mg) disp. scored: 0.1** | **ABC/3TC (120/60 mg) disp. scored: 0.1** | **ABC/3TC (600/300 mg): 0.25** | **ABC/3TC (600/300 mg): 0.25** |
| | **Daily cost($)** | **0.25** | **0.3** | **0.25** | **0.25** |
| **Zidovudine/ Lamivudine ZDV/3TC** | Daily dose | 300/150 mg | 360/180 mg | 600/300mg | 600/300mg |
| | Tablets taken | 3*60/30 mg ZDV/3TC (AM) + 2*60/30 mg ZDV/3TC[2] (PM) | 3*60/30 mg ZDV/3TC (AM) + 3*60/30 mg ZDV/3TC[2] (PM) | 1*300/150 mg ZDV/3TC (AM) + 1*300/150 mg ZDV/3TC (PM) | 1*300/150 mg ZDV/3TC (AM) + 1*300/150 mg ZDV/3TC (PM) |
| | **Unit cost($)** | **ZDV/3TC (60/30 mg) disp. scored: 0.03** | **ZDV /3TC (60/30 mg) disp. scored: 0.03** | **ZDV /3TC (300/150 mg): 0.09** | **ZDV /3TC (300/150 mg): 0.09** |
| | **Daily cost($)** | **0.15** | **0.18** | **0.18** | **0.18** |
| **Tenofovir alafenamide/ Emtricitabine TAF/FTC** | Daily dose | 120/15 mg | 120/15 mg | 200/25 mg | 200/25 mg |
| | Tablets taken | 1*120/15 mg FTC/TAF | 1*120/15 mg FTC/TAF | 1*200/25 mg FTC/TAF | 1*200/25 mg FTC/TAF |
| | **Unit cost($)** | **TAF/FTC/DTG (25/200/50 mg): 0.17** | **TAF/FTC/DTG (25/200/50 mg): 0.17** | **TAF/FTC/DTG (25/200/50 mg): 0.17** | **TAF/FTC/DTG (25/200/50 mg): 0.17** |
| | **Daily cost($)** | **0.102** | **0.102** | **0.17** | **0.17** |

Notes: ART costs were calculated using data on usage from CHAPAS-4 trial and unit costs of different ART drugs from Clinton Health Access Initiative (CHAI) where available (2022). Where data was unavailable, trial sites provided information on costs. We used proportions (calculated based on dosage) of the published adult drug costs for the costs of the paediatric drugs 120/15 mg FTC/TAF. ABC: abcavir, 3TC: lamivudine, ZDV: zidovudine, TAF: tenofovir alafenamide, FTC: emtricitabine, DTG: dolutegravir

1. or 5 x ABC/3TC 120/60mg;

2. or 2.5+2.5;

3. Tablets taken: once daily if not stated, or twice daily (morning **AM** and afternoon **PM**)

Table CES2: Costs of resources used

| | Uganda | | Zambia | | Zimbabwe | |
|---|---|---|---|---|---|---|
| | 2022 price in US$ (adjusted for local inflation) | Source | 2022 price in US$ (adjusted for local inflation) | Source | 2022 price in US$ (adjusted for Zambia's inflation as a proxy) | Source |
| Hospital overnight: Teaching hospital | 4.81 | WHO Choice (2011) | 8.83 | WHO Choice (2011) | 3.93 | WHO Choice (2011) |
| Outpatient attendances: Health Centre (no beds) | 0.97 | | 1.78 | | 0.75 | |
| Outpatient attendances: Secondary-level hospital | 1.42 | | 2.60 | | 1.09 | |

Notes: 1. Unit costs of outpatient attendances in a secondary-level hospital (the highest level) were used to cost scheduled and unscheduled visits, unit costs of hospital overnight in a teaching hospital (the highest level) were used to cost hospital stays, and unit costs of outpatient attendances in a health centre (no beds) were used to calculated the costs of visiting a local clinic or healthcare workers; 2. All unit costs were checked with the CHAPAS-4 trial sites to confirm that they were reasonable estimates of the actual costs; 3. Unit cost information collected before 2022 were adjusted for local inflation to get the 2022 price (the unit cost was reported in US dollar in WHO Choice (2011).

Table CES3. Resource Use by NRTI backbone randomization groups.

| NRTI Backbone Randomization Groups | SOC | | TAF/FTC | |
|---|---|---|---|---|
| | Mean | SD | Mean | SD |
| *ART use* | | | | |
| Average duration of four anchor drugs (days) | 662.30 | 57.79 | 666.07 | 47.93 |
| LPV/r duration (days) | 651.90 | 85.91 | 652.91 | 85.07 |
| As % of the 96-week trial on LPV/r | 97.01% | 12.78% | 97.16% | 12.66% |
| N (as % of the 919 children) | 115 (12.51%) | | 112 (12.19%) | |
| ATV/r duration (days) | 655.89 | 55.37 | 664.06 | 29.46 |
| As % of the 96-week trial on ATV/r | 97.60% | 8.24% | 98.82% | 4.38% |
| N (as % of the 919 children) | 115 (12.51%) | | 116 (12.62%) | |
| DRV/r duration (days) | 658.16 | 69.81 | 659.96 | 67.26 |
| As % of the 96-week trial on DRV/r | 97.94% | 10.39% | 98.21% | 10.01% |
| N (as % of the 919 children) | 114 (12.4%) | | 118 (12.84%) | |
| DTG duration (days) | 667.74 | 23.38 | 649.66 | 99.44 |
| As % of the 96-week trial on DTG | 99.37% | 3.48% | 96.68% | 14.80% |
| N (as % of the 919 children) | 117 (12.73%) | | 112 (12.19%) | |
| Average duration of NRTI backbone drugs (days) | 661.78 | 56.83 | 663.79 | 54.49 |
| Average duration of other ART drugs (days) | 0.24 | 5.22 | 2.80 | 42.36 |
| *Other health care use* | | | | |
| Scheduled clinic visits | 10.76 | 0.87 | 10.82 | 0.67 |
| Unscheduled clinic visits | 1.22 | 2.27 | 0.97 | 1.85 |
| Hospital stays (days) | 0.24 | 2.40 | 0.19 | 1.49 |
| Visits to a local clinic or healthcare workers | 0.17 | 0.50 | 0.27 | 1.18 |
| N (as % of the 919 children) | 461 (50.16%) | | 458 (49.84 %) | |

Notes: 1. The days in four anchor drugs were calculated conditional on patients being allocated to the specific anchor drugs at randomization; 2. We only calculated the resources used when patients were on second-line treatment; 3. Four anchor drugs included LPV/r, ATV/r, DRV/r, DTG; 4. NRTI backbone drugs included TAF/FTC and SoC (ABC/3TC or ZDV/3TC); 5. Other ART drugs included TDF/3TC/DTG (300/300/50 mg) and TDF/3TC. NRTI: Nucleos(t)ide reverse transcriptase inhibitor, SOC: standard-of-care, TAF: tenofovir alafenamide, FTC: emtricitabine, SD: standard deviation, ART: antiretroviral therapy, LPV/r: ritonavir-boosted lopinavir, ATV/r: ritonavir-boosted atazanavir, DRV/r: ritonavir-boosted darunavir, DTG: dolutegravir.

Table CES4. Costs and QALYs by NRTI backbone randomization groups

| NRTI backbone randomization groups | SOC | | TAF/FTC | |
|---|---|---|---|---|
| Costs | Mean | SD | Mean | SD |
| *ART costs* | | | | |
| Anchor drugs costs | 236.01 | 131.74 | 244.79 | 132.94 |
| DTG cost | 13.29 | 23.53 | 12.19 | 21.81 |
| ATV/r cost | 59.69 | 105.63 | 61.79 | 107.67 |
| LPV/r cost | 66.55 | 120.08 | 68.46 | 122.96 |
| DRV/r cost | 93.31 | 165.43 | 99.05 | 170.73 |
| NRTI backbone drugs costs | 140.43 | 32.37 | 98.91 | 21.70 |
| Other ART drug costs | 0.03 | 0.68 | 0.30 | 4.48 |
| **Total ART costs** | **371.39** | **136.78** | **339.35** | **137.49** |
| *Other health care costs* | | | | |
| Scheduled visits costs | 14.94 | 5.52 | 15.09 | 5.53 |
| Unscheduled visits costs | 1.59 | 2.84 | 1.28 | 2.32 |
| Hospital stay costs | 1.30 | 11.07 | 0.94 | 6.89 |
| Cost of visiting a local clinic or healthcare workers | 0.23 | 0.75 | 0.31 | 1.10 |
| **Total other health care costs** | **18.05** | **14.06** | **17.61** | **10.20** |
| **Total health care costs** | **389.35** | **136.95** | **356.97** | **138.52** |
| **QALYs** | **1.8035** | **0.0285** | **1.7992** | **0.0873** |
| N (as % of the 919 children) | 461 (50.16%) | | 458 (49.84 %) | |

Notes: 1. All the costs were in 2022 US dollar; 2. The discount factor for costs and QALYs occurring in in the second year of the study was 3% per annum; 3. Anchor drugs included DTG , ATV/r, LPV/r, DRV/r; 4. NRTI backbone drugs included TAF/FTC and SoC (ABC/3TC or ZDV/3TC); 5. Other ART drugs included TDF/3TC/DTG (300/300/50 mg) and TDF/3TC; 6. QALYs were captured by EQ-5D index over 96 weeks using area-under-the-curve approach. Only EQ-5D indices reported in the scheduled visits were used to calculate QALYs. Missing baseline indices were imputed using the sample average index, while other missing indices were imputed using the mean of the indices before and after the missing index of the same individual. NRTI: Nucleos(t)ide reverse transcriptase inhibitor, SOC: standard-of-care, TAF: tenofovir alafenamide, FTC: emtricitabine, SD: standard deviation, ART: antiretroviral therapy, LPV/r: ritonavir-boosted lopinavir, ATV/r: ritonavir-boosted atazanavir, DRV/r: ritonavir-boosted darunavir, DTG: dolutegravir, QALY: quality-adjusted life year

Table CES5. Costs Analysis of SOC versus TAF/FTC

| Comparators | Total Costs | Incremental cost | | | | Probability of being least costly |
|---|---|---|---|---|---|---|
| | mean | mean | SE | 95% CI | | |
| SOC | 391.61 | | | | | 0% |
| TAF/FTC | 353.92 | -37.68 | 3.39 | -44.32 | -31.05 | 100% |
| N=919 | | | | | | |

Notes: 1. All the costs were in 2022 US dollar and a discount rate of 3% per annum was applied to costs incurred for participants in their second year of the study; 2. The model controlled for stratification factors of the six sites and the NRTI backbone drugs they failed on in the first line treatment; 3. Probabilistic sensitivity analyses were conducted to estimate the probability of each intervention being least costly using the threshold of \$500 per QALY implied by the decisions that had been previously made on the ART drugs for HIV. SOC: standard-of-care, TAF: tenofovir alafenamide, FTC: emtricitabine, SE: standard error, CI: confidence interval.

Table CES6. Cost-effectiveness of SOC versus TAF/FTC

| Comparators | Mean Cost | Mean QALYs | Incremental cost | | | | Incremental QALYs | | | | ICER | INHB (λ=100) | Prob of CE | INHB (λ=300) | Prob of CE | INHB (λ=500) | Prob of CE |
|---|---|---|---|---|---|---|---|---|---|---|---|---|---|---|---|---|---|
| | mean | mean | mean | SE | 95% CI | | mean | SE | 95% CI | | | | | | | | |
| SOC | 391.61 | 1.8037 | | | | | | | | | | | 0% | | 0% | | 0% |
| TAF/FTC | 353.92 | 1.7990 | -37.68 | 3.39 | -44.32 | -31.05 | -0.0048 | 0.0041 | -0.0129 | 0.0033 | 7860.87 | 0.3728 | 100% | 0.1211 | 100% | 0.0707 | 100% |
| N=919 | | | | | | | | | | | | | | | | | |

Notes: 1. All the costs were in 2022 US dollar and the discount factor for costs and QALYs occurring in in the second year of the study was 3% per annum; 2. Generalized linear model with a gamma distribution of the dependent variable and an identity link function was used to predict the mean total costs and estimate the incremental costs, controlling for the stratification factors; 3. Generalized linear model with a gaussian (normal) distribution of the dependent variable and an identity link function (equivalent to ordinary least squares) was used to predict the mean QALYs and estimate the incremental QALYs, controlling for the stratification factors and the baseline EQ5D indices; 4. Cost-effectiveness threshold of $500 per QALY was used to calculate ICER based on the decisions that had been previously made on the ART drugs for HIV; 5. cost-effectiveness thresholds of $100 and $300 per QALY were also used when calculating INHB for scenario analyses, and probabilistic sensitivity analyses were conducted to estimate the probability of each intervention being cost-effective (CE) using the three thresholds of $500, $300 and $100 per QALY. QALY: quality-adjusted life year, INHB: incremental net health benefit, CE: cost effectiveness.
